# A Genetically Driven Immunologic Mechanism Underlying the Link between EBV and Multiple Sclerosis

**DOI:** 10.64898/2025.12.11.25342083

**Authors:** Yoshiaki Yasumizu, Namkwon Kim, Cyprien A. Rivier, Jeonghyeon Moon, Shohei Kojima, Heng-Le Chen, Nicholas Buitrago-Pocasangre, Andrew Silberfeld, Elizabeth Quinn, Stephen Vaughn, Annalisa Morgan, Shufan Huo, Tomokazu S. Sumida, Kazuyoshi Ishigaki, Erin E. Longbrake, Guido J. Falcone, David A. Hafler

## Abstract

Epstein–Barr virus (EBV) is implicated as a trigger of multiple sclerosis (MS), yet the host genetic mechanisms linking EBV activity to MS is unknown. We performed a cross-ancestry genome-wide association study of EBV DNA positivity (N=617,186), identifying 39 susceptibility risk loci that significantly overlapped with MS risk genes (p=1.3×10^-12^). Using our single-cell method for EBV detection, we identified 1,069 EBV-infected B cells across 38 individuals. EBV was predominantly in the latent phase and B cell receptor analysis revealed that memory B cell differentiation is altered by infection. Notably, EBV-infected switched memory and atypical B cells upregulated cytokines and costimulatory signals that influence T cell activation, and B cell signaling pathway. Finally, EBV-infected memory B cells upregulated risk genes from both the EBV and MS, suggesting that EBV-infected B cells link T cell modulation to activation of MS susceptibility pathways. These findings define a genetically driven immunologic mechanism underlying MS.

## Introduction

Multiple sclerosis (MS) is the most frequent central nervous system (CNS) disease of young adults. The disease most commonly begins with an autoimmune, relapsing remitting phase mediated by circulating autoreactive T cells^1^ that migrate to the central nervous system (CNS) and induce a marked inflammatory response both in the white matter and cortical grey matter. A secondary, neurodegenerative disease course occurs in a significant number of patients, presumably triggered by the initial T cell-mediated autoimmune process. As with other autoimmune disorders, MS is triggered by environmental interactions in genetically susceptible individuals. Indeed, we have identified 233 common genome loci associated with disease risk and one variant associated with disease progression.^2–4^ The majority of these map to the immune system, targeting T cells, regulatory T cells, and, in particular, B cells.^2,5,6^ As a result, MS patients exhibit a spectrum of immunopathology, including a skewing of T cells toward pro-inflammatory Th1 and Th17 programs,^6–8^ loss of regulatory T-cell function with secretion of IFNγ,^9,10^ and activation of B cells.^11^

Compelling epidemiologic data implicates Epstein–Barr virus (EBV) as the initiating event breaking immune tolerance in genetically susceptible individuals. A large longitudinal study of U.S. military personnel reported that EBV infection occurs before MS development^12^, indicating that EBV is a necessary causal factor for MS. Serum neurofilament light chain, a marker of axon injury, begins to rise only after EBV seroconversion. Moreover, MS patients exhibit higher EBV activity,^13–15^ increased frequencies of EBV-reactive T cells^16,17^, and EBV-derived antigens that cross-react with myelin antigens ^18–20^. EBV primarily infects B cells, directly influencing their function^21^, and we have observed distinctive alterations in B-cell phenotypes in MS (*manuscript in preparation*). Importantly, B-cell depletion therapy is a highly effective treatment for the early relapsing remitting, autoimmune phase of MS,^22^ underscoring the central role of B cells in disease pathogenesis.

EBV plays a causal role in several other human diseases. Epidemiologic and immunologic evidence links EBV to non-Hodgkin lymphoma, including Burkitt lymphoma, nasopharyngeal carcinoma, gastric carcinoma, and, as discussed above, to MS, rheumatoid arthritis (RA), and systemic lupus erythematosus (SLE).^23^ Although EBV infection is ubiquitous, affecting more than 90% of the global population, its clinical manifestations vary widely. In most individuals, EBV infection remains asymptomatic or latent, whereas a subset of subjects develops infectious mononucleosis or chronic active EBV infection. Moreover, infectious mononucleosis is not only a symptomatic form of EBV infection but also a strong epidemiological risk factor for MS.^24^ However, few individuals develop malignancies or autoimmune diseases after EBV infection. This variability suggests that the host’s ability to control EBV reactivation differs among individuals, potentially due to underlying genetic factors.^25^

EBV activity can be assessed through two major biomarkers: anti-EBV antibodies and EBV DNA levels. Immunoglobulin (Ig) G targeting EBV antigens reflects prior exposure. Over 90% of healthy adults and nearly 100% of patients with MS are seropositive.^15,26^ In contrast, circulating EBV DNA provides a more direct readout of viral persistence, reactivation burden, or reservoir dynamics; blood EBV DNA has been reported to be elevated in patients with MS and SLE.^14,27^ Circulating EBV DNA is also clinically useful for nasopharyngeal carcinoma screening.^28^ These complementary biomarkers therefore capture distinct aspects of EBV biology and may reveal different dimensions of host genetic control.

Genome-wide association studies (GWAS) have enabled the identification of host genetic variants influencing susceptibility to infection-related traits and pathogen burden. For example, analysis of HIV viral load overcame the limited resolution of infection-status GWAS and identified key loci in the HLA and *CCR5* regions.^29^ Recent advances in sequencing technologies now allow simultaneous quantification of host and viral genomes from the same dataset. Our group previously developed *VIRTUS*,^30^ a pipeline for detecting viral transcripts from RNA-seq data, and EBV detection from bulk RNA-seq has also been reported by others.^31^ Similar WGS-based approaches have recently been applied to the human blood virome, including a GWAS of human herpesvirus 6 (HHV-6) viral load that revealed integration sites and links to SLE disease activity.^32^ For EBV, prior host-genetic studies have largely focused on serologic antibody titers,^33^ which capture infection history rather than DNA-based measures of viral activity. More recently, population-scale WGS studies have demonstrated that EBV-derived reads can be quantified in large biobanks and that host genetic determinants, particularly within the MHC, contribute to variation in EBV DNA detection and persistence. These studies establish the feasibility of EBV DNA GWAS at a biobank scale while motivating the need to define how these signals extend across ancestry groups and connect to disease-relevant cellular phenotypes. ^34–36^

Here, we analyzed 617,186 participants from the All of Us Research Program^37^ and the UK Biobank^38^ using harmonized viral-detection and quality-control pipelines across ancestries and performed a cross-ancestry meta-analysis of EBV DNA positivity and viral load. We extend this framework by jointly analyzing EBV DNA positivity and viral load, integrating phenome-wide association studies (PheWAS), polygenic risk score (PRS) analyses, Mendelian-randomization (MR)-based analyses, and HLA and *ERAP2* interpretation to examine the shared host-genetic architecture linking EBV activity to human disease (Fig. S1). Combining these population-scale analyses with single-cell RNA-seq (scRNAseq) and spatial transcriptomic analyses, we identified specific cell types and anatomical niches involved in EBV control. Notably, using *VIRTUS3*, we successfully identified EBV-infected cells from people with MS and characterized their genetic effects. Together, these findings provide a multi-ancestry genetic and cellular framework for host control of EBV activity. By integrating cross-ancestry host genetics with single-cell analysis of EBV-infected B cells in people with MS, our study links population-scale EBV DNA signals to disease-relevant cellular states and offers a mechanistic context for EBV-associated autoimmunity, elucidating a potential molecular mechanism underlying the cause of MS.

## Results

### EBV DNA detection from WGS

To detect EBV reads in WGS datasets, we developed an analysis pipeline that filters and counts reads mapped to the EBV decoy sequence in the human reference genome. We applied our pipeline to the diverse cohort of study participants enrolled in the All of Us Research Program. After quality control, 409,331 individuals were included. DNA information was obtained from two sources: whole blood (*N*=359,796) and saliva (*N*=48,086). EBV DNA was detected in 20.8% of blood samples and 52.7% of saliva samples. Age, sex, and ancestry were significantly associated with EBV positivity. In both blood and saliva, higher age was strongly associated with higher detection rates (Fig. 1A, blood: *OR*=1.16 per 10 years, 95% *CI*=1.15–1.16, *p*<1×10^-100^; saliva: *OR*=1.10 per 10 years, *95% CI*=1.09–1.11, *p*<1×10^-65^). Female sex was associated with a lower rate of EBV positivity compared with males (Fig. 1A, blood: *OR*=0.83, *95% CI*=0.81–0.84, *p*<1×10^-100^; saliva: *OR*=0.78, *95% CI*=0.75–0.81, *p*<1×10^-35^).

**Figure 1.**
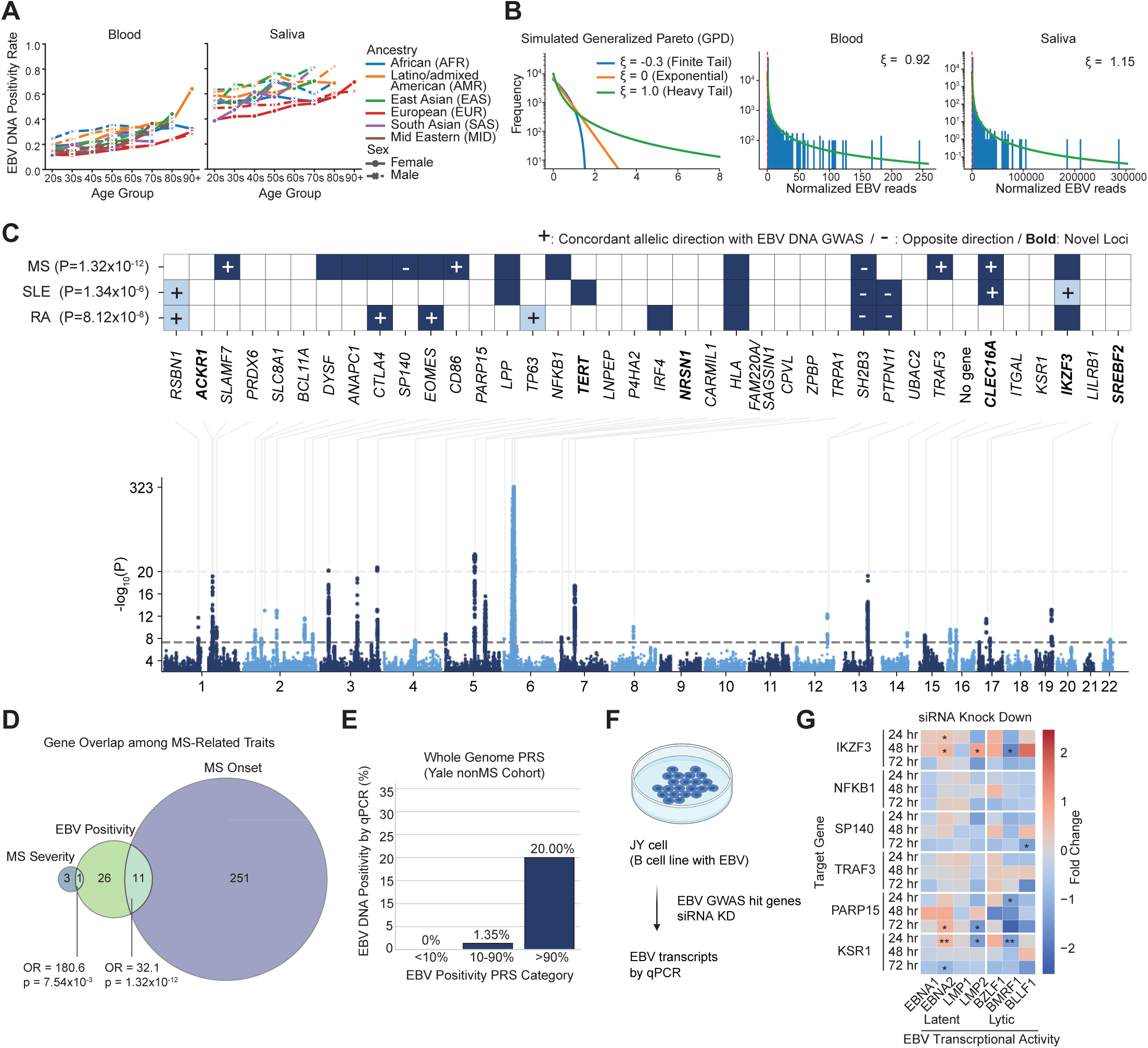
EBV DNA GWAS reveals genetic architecture of EBV regulation (A) EBV DNA positivity across predicted ancestors, gender, and age in blood (left) and saliva (right). AFR, African ancestry; AMR, Latino/admixed American ancestry; EAS, East Asian ancestry; EUR, European ancestry; SAS, South Asian ancestry (B) Distribution of EBV DNA fitted with Generalized Pareto distribution (GPD). Left: Simulated examples of GPD probability density functions illustrating the effect of the shape parameter ξ on the tail behavior: finite tail (ξ < 0), exponential tail (ξ = 0), and heavy tail (ξ > 0). Middle and Right: Histograms of normalized EBV DNA reads in blood (middle) and saliva (right). Orange curves represent GPD fits above the 95th percentile threshold (red dashed line). Estimated ξ values are shown for each sample type. (C) Manhattan plots showing genome-wide association statistics of EBV DNA presence. The y-axis shows-log_10_P of each variant, and lead SNPs and annotated genes were highlighted. Heatmap shows the genes previously reported in MS,^2,4,119^ SLE,^120^ and RA^121^ GWAS. The dashed line indicates a genome-wide significant level (*p*=5.0×10^-8^). Symbols indicate effect direction relative to the EBV GWAS (+ concordant, − discordant). Directional symbols are displayed only for loci reaching *P*<5×10⁻^5^ in the respective MS/SLE/RA GWAS, while nonsignificant or unavailable associations are shown as blank cells. Genes not reported in the GWASs with *P*<5×10⁻^5^ in the most significant loci of EBV DNA positivity GWAS were highlighted in light blue. Details are also available in Table S4. (D) Overlap of genes highlighted in MS onset GWAS,^2^ MS severity GWAS^4^ and EBV positivity GWAS. Statistical significance was evaluated using a two-tailed Fisher’s exact test on a 2×2 contingency table, with all protein-coding genes (Gencode v47) used as the background (C and D). (E) EBV DNA positivity measured by qPCR across categories of the EBV DNA positivity PRS in an independent Yale cohort of European-ancestry individuals without MS (N = 94; 3 EBV DNA^+^ cases). (F) Schematic overview of the experimental design. JY cells, an EBV-positive B-cell line, were subjected to siRNA-mediated knockdown of host genes prioritized from the EBV DNA GWAS, and EBV transcripts were quantified by qPCR. (G) Heatmap showing changes in EBV transcript levels after knockdown of the indicated host genes at 24, 48, and 72 hours. Columns represent latent-associated (EBNA1, EBNA2, LMP1, LMP2) and lytic-associated (BZLF1, BMRF1, BLLF1) EBV transcripts. Color indicates fold change in transcript expression, with red indicating increased expression and blue indicating decreased expression. Asterisks indicate statistically significant changes tested by the Welch two-sample t-test. *: p < 0.05, **: p < 0.01, ***: p < 0.001

When examining ancestry, Europeans had the lowest EBV positivity, whereas all other ancestries showed significantly higher odds ratios of EBV presence. In blood, African (*OR*=2.17, *p*<1×10^-100^), Latino/admixed American (*OR*=2.02, *p*<1×10^-100^), East Asian (*OR*=1.47, *p*=9.2×10^⁻43^), Middle Eastern (*OR*=1.31, *p*=1.2×10⁻^4^), and South Asians (*OR*=1.29, *p*=6.1×10⁻^11^) ancestries had significantly higher odds ratios relative to Europeans (Fig. S2A). In saliva, African (*OR*=1.61, *p*=4.1×10⁻^65^), Latino/admixed American (*OR*=2.02, *p*<1×10^-100^), East Asian (*OR*=1.94, *p*=5.8×10⁻^32^) and South Asian ancestries also showed increased odds (*OR*=1.42, *p*=1.4×10⁻^5^) (Fig. S2A). Middle Eastern ancestry showed a trend for higher EBV positivity in saliva but did not reach statistical significance (*OR*=1.32, *p*=0.078).

Next, we examined the distribution of EBV DNA load. When normalized by the number of reads mapped to the human genome, the EBV DNA load was on average 3,826-fold higher in saliva compared to blood (Fig. 1B). Moreover, a subset of individuals carried viral loads that far exceeded the population average. Indeed, in blood, the top 1% of samples accounted for 64% of the total EBV DNA load, with the top 5% and 10% contributing 83% and 91%, respectively. In saliva, the skew was even more pronounced: the top 1% accounted for 80%, the top 5% for 94%, and the top 10% for 97%. To statistically evaluate this trend, we approximated the distribution with a generalized Pareto distribution (GPD). A positive shape parameter (ξ) indicates a heavy-tailed distribution (Fig. 1B). In blood, ξ was estimated as 0.92 (95% *CI*:0.86–0.98) at the 95th percentile threshold, while in saliva, ξ was even larger at 1.15 (95% *CI*:1.04–1.25). Varying the threshold from the 90th to 99th percentiles yielded consistently positive ξ values, confirming the robustness of these results. These findings demonstrate that the distribution of EBV DNA load is heavy-tailed in both blood and saliva, suggesting the presence of a small number of super-carriers who contribute disproportionately to the overall viral burden.

To comprehensively investigate diseases co-occurring with EBV infection, we conducted a Phenome-wide Association Study (PheWAS). Using European-ancestry participants from All of Us, we performed a PheWAS of EBV DNA positivity and viral load (Figs. S2B, C, Supplementary Tables 1 and 2). As recently reported,^35^ we confirmed that both EBV DNA positivity and viral load showed strong associations with immunosuppressive conditions such as HIV infection and post-transplant immunosuppression, as well as with tobacco use and chronic obstructive pulmonary disease (COPD).^39^ Furthermore, the analysis identified a significant association with infectious mononucleosis, a manifestation of primary EBV infection in adolescents. Unexpectedly, neither MS nor SLE emerged as positively associated traits (MS: EBV positivity, *OR*=0.83, *FDR*=0.034; viral load, *FDR*=0.78. SLE: EBV positivity, *FDR*=0.27; viral load, *FDR*=0.96). In addition to confirming known associations with rheumatoid arthritis, we observed correlations with encephalitis, encephalopathy, and polyneuropathies, suggesting links to other inflammatory neurological diseases. Although more traits were significantly associated with EBV DNA positivity (*FDR*<0.05, 475 traits) than with EBV viral load (270 traits), the viral load analysis specifically highlighted associations with virus-related malignancies such as non-Hodgkin lymphoma and T cell lymphoma, as well as myopathy. The PheWAS results not only reproduced well-established associations between EBV and related diseases, validating our analytical pipeline, but also revealed broader links to autoimmune and inflammatory disorders, underscoring the extensive disease spectrum influenced by EBV infection.

### Multi-Ancestry genome-wide association study of EBV DNA

We applied a similar pipeline to the UK Biobank, in which WGS was performed using DNA derived from whole blood. EBV-derived DNA was detected in 15.5% of UK Biobank samples. We then performed a cross-ancestry GWAS combining UK Biobank samples with blood-derived samples from the All of Us cohort. We selected 5,020,877 SNPs with a minor allele frequency (MAF) of ≥1% in All of Us (Methods). Association tests were performed using Regenie^40^ in all ancestry groups from the All of Us Research Program and in European participants from the UK Biobank. A European meta-analysis was conducted using METAL^41^ (N=493,312), and a cross-ancestry meta-analysis (European from UK Biobank and European, African, and Latino/Admixed American from All of Us) was performed using MR-MEGA^42^ (N=617,408) (Supplementary Table 3). In the cross-ancestry analysis, we identified 39 genome-wide significant loci associated with EBV positivity and 16 loci associated with viral load (Figs. 1C, S3A, Supplementary Tables 4-6). Using LD score regression (LDSC),^43^ the SNP-based heritability (h^2^) of EBV positivity was estimated at 1.77% (*SE*=0.22%), while that of viral load was 1.24% (*SE*=0.18%). There was a positive genetic correlation between EBV positivity and EBV viral load (*rg*=0.71, *p*=4.08×10^-24^). We observed a slight inflation of test statistics in the cross-ancestry meta-analysis, particularly for the binary trait (Fig. S3B, EBV positivity, λ*_GC_*=1.15; viral load, λ*_GC_*=1.07). Importantly, the λ*_GC_* values from each individual GWAS were well controlled (λ*_GC_*<1.08 in all cases, Supplementary Table 3). We evaluated whether the MHC contributed to the inflation; removing this region yielded comparable values (EBV positivity, λ*_GC_*=1.14; viral load, λ*_GC_*=1.06). Of note, in the LDSC analysis, intercepts were low (EBV positivity, *intercept*=1.04; viral load, *intercept*=1.03), indicating that the observed inflation is likely due to polygenicity rather than residual confounding.

Next, we examined the lead SNPs within each locus in detail. Outside of the HLA region, approximately half of the associated variants were intronic (Supplementary Table 4, positivity: 63.1%; viral load: 53.3%), and roughly half were located in cis-eQTLs (Supplementary Table 4, positivity:55%; viral load: 60%). The majority of these loci (Supplementary Table 4, positivity: 65.8%; viral load: 60%) have been previously implicated in immune-related traits, including MS, SLE, RA, IgM levels, and blood cell counts in prior GWAS (Fig. 1C). Remarkably, there was a significant overlap with genetic variants associated with MS risk (Fig. 1D, *OR*=32.1, *p*=1.3×10^-12^) and severity (Fig. 1D, *OR*=180.6, *p*=7.54×10^-3^) including MS susceptibility alleles, *HLA-DRB1*\*04:04, *HLA-A*\*02:01 and to a lesser extent, *HLA-DRB*1*15:01. Specifically, among the top hits, 29.7% of the EBV positivity-associated genes and 33% of the viral load-associated genes overlapped with genes reported in our MS GWAS, including *DYSF*, which has been associated with MS severity^4^ (Figs. 1C and S4A). When we examined the direction of effects for EBV DNA positivity across autoimmune diseases, we found that a number genetic variants, including *SLAMF7*, *CTLA4*, *EOMES*, *CD86*, *TP63*, *TRAF3*, *CLEC16A*, *RSBN1,* and *IKZF3* showed concordant effects, with increased EBV DNA positivity associated with increased disease risk, while *SP140*, *SH2B3*, and *PTPN11* exhibited discordant directions of effect with less EBV DNA presence (Fig. 1C). The genes highlighted by the GWAS provided biological insights into the regulation of EBV activity. For example, *TERT*, encoding the catalytic subunit of telomerase, emerged as one of the top hits. Indeed, the top variant within this locus, rs7726159, is associated with telomere length^44^ (Supplementary Table 5, Fig. S4B). Given that EBV is known to drive telomerase regulation to promote immortalization of infected cells,^45^ this finding may highlight a mechanistic link between EBV replication and telomere biology. We also observed strong support for biologically plausible genes, including *IFI16*, a nuclear DNA sensor that restricts herpesvirus reactivation, and *SP140*, a PML-nuclear body component involved in antiviral chromatin regulation by repressing type I IFN responses (Fig. S4C).^46–48^

Stratified LD score regression^49^ revealed that genetic heritability for both EBV DNA positivity and EBV DNA load was concentrated in transcriptionally active genomic regions, particularly at transcription start sites (TSS; enrichment=23.4±4.1 [positivity], 31.3±5.3 [load]), FANTOM5 enhancers (19.9±10.3 [positivity], 32.7±13.8 [load]), and coding regions (17.2±5.2 [positivity], 17.0±6.3 [load]), indicating that genetic variants influencing EBV control tend to localize to regulatory and coding elements with high transcriptional activity (Fig. S4D). Pathway enrichment analysis revealed that EBV susceptibility loci were primarily concentrated in immune response–related pathways (Fig. S4E, Supplementary Tables 7 and 8). Both EBV positivity and EBV DNA load showed significant enrichment for T cell activation, CD8^+^ αβ T cell activation, and T cell proliferation, reflecting a shared involvement of cytotoxic T cell–mediated immune responses. Cell cycle G1/S phase transition was specifically enriched for EBV positivity, while EBV DNA load additionally involved epithelial-associated pathways such as positive regulation of keratinocyte proliferation and multiciliated epithelial cell differentiation, indicating potential contributions of epithelial cell proliferation and differentiation to viral load regulation.

To assess the effects of GWAS-prioritized host genes on EBV transcriptional activity, we performed siRNA-mediated knockdown of candidate genes and analyzed EBV transcripts (Fig. 1F). Knockdown of EBV DNA GWAS-prioritized host genes altered EBV transcriptional activity and, at 72 hours, was associated with a shift toward reduced EBV gene expression (Figs. 1G and S6). This was more pronounced for lytic-associated transcripts than for latent-associated transcripts. Consistent with this pattern, for example, *SP140* knockdown reduced *BLLF1* expression, whereas *PARP15* knockdown decreased *LMP2* and *BMRF1*. However, these effects were not uniform, including a transient increase in latent transcripts after *IKZF3* knockdown and increased *EBNA2* expression at 72 hours after *PARP15* knockdown. Together, these findings suggest that EBV DNA GWAS hit genes contribute to the regulation of EBV transcription, but their effects are not simply binary; rather, they act in a transcript-selective and time-dependent manner, with a broader repressive trend emerging at later time points.

To further test the causal relationship between EBV and autoimmune diseases, we performed bidirectional Mendelian randomization using 27 independent SNPs associated with EBV DNA positivity (Fig. S5A, Supplementary Tables 9). Genetically predicted EBV DNA positivity was associated with increased risk of MS (OR=1.45, 95% CI=1.18–1.80), RA (OR=1.27, 95% CI=1.05–1.54), and SLE (OR=1.44, 95% CI=1.08–1.91), but not ischemic stroke, used as a negative control. These effects were predominantly driven by MHC variants, and reverse MR analyses did not support an effect of autoimmune disease liability on EBV DNA levels. Together, these results support a unidirectional contribution of genetically determined EBV activity to autoimmune disease risk, largely mediated by MHC variation.

To investigate whether the polygenic architecture of EBV activity contributes to autoimmune disease risk, we constructed PRSs from the EBV DNA GWAS in the European ancestry subset of the All of Us cohort and tested them in European ancestry participants in the UK Biobank (N=487,181; 2,423 MS cases; Fig. S5B, Supplementary Tables 10). A one–standard deviation increase in the EBV DNA positivity PRS was associated with increased risk of MS (OR=1.11, 95% CI=1.07–1.16, p<0.0001), with individuals in the top PRS decile showing higher MS prevalence than those in the bottom decile (0.68% vs 0.52%; OR=1.33, 95% CI=1.13–1.57). Partitioning the PRS revealed that this association was largely driven by variants within the MHC region, whereas non-MHC variants showed no significant effect. Together, these results indicate that host genetic variation shapes EBV activity, with a substantial contribution from immune regulatory loci and the MHC region, linking genetically determined EBV control to autoimmune disease susceptibility.

### Polygenic risk scoring and EBV DNA measured by quantitative PCR (qPCR)

To evaluate whether identified genome-wide significant risk loci for EBV DNA capture biologically meaningful variation in viral load, we assessed PRSs in an independent cohort with qPCR-based measurement of EBV DNA. We calculated PRSs based on risk variants identified in the meta-analysis of European ancestry using PRScs.^50^ We evaluated association with EBV DNA positivity measured by qPCR in an independent Yale cohort of European ancestry (Fig. 1E, S5C, and Supplementary Tables 11; *N*=94; mean age 40.2±14.7 years; 71% female). EBV DNA positivity was 2/10 (20%) in the top decile, 1/74 (1.4%) in the middle group, and 0/10 (0%) in the bottom decile for both positivity and viral load PRS (Ordinal trend tests positivity: *p*=0.024; viral load: *p*=0.016). Consistently, one standard deviation increase in the PRS was associated with a higher EBV DNA load after adjustment (log10 EBNA1 copies/mL) (positivity: β=0.21, *SE*=0.076, *p*=0.006; load: β*=0.26*, *SE*=0.075, *p*=0.0009). When we stratified variants by their location inside or outside the MHC region, only the PRSs derived from the MHC region showed significant associations with qPCR-confirmed EBV DNA positivity and load, while PRSs constructed from variants outside the MHC region showed no significant associations. Taken together, these findings demonstrate that qPCR-measured EBV DNA levels in our independent cohort closely track with the polygenic architecture identified by our EBV DNA GWAS, supporting the validity and biological relevance of the GWAS results.

### Antigen presentation by HLA and EBV activity

The strongest associations in both GWASs were located within the HLA locus. In addition, a secondary genome-wide significant signal was identified at rs27300, a variant residing within a linkage disequilibrium block encompassing the *LNPEP* and *ERAP1/2* genes (Figs. 2A and S6A). This variant exhibited a significant eQTL effect on *ERAP2* expression in lymphoblastoid cell lines (LCLs) (*p*=2.33×10^-118^, Fig. 2B), but not with *LNPEP* (*p*=7.07×10^-6^, Fig. 2B). *ERAP2* encodes an aminopeptidase essential for peptide trimming selectively during antigen presentation via MHC class I molecules. This allelic directionality provides additional support for a functional role of *ERAP2* in EBV control. The *ERAP2*-increasing C allele corresponds to the protective direction in the EBV GWAS, suggesting that more efficient peptide trimming enhances antigen presentation and improves the clearance of EBV-infected cells.

**Figure 2.**
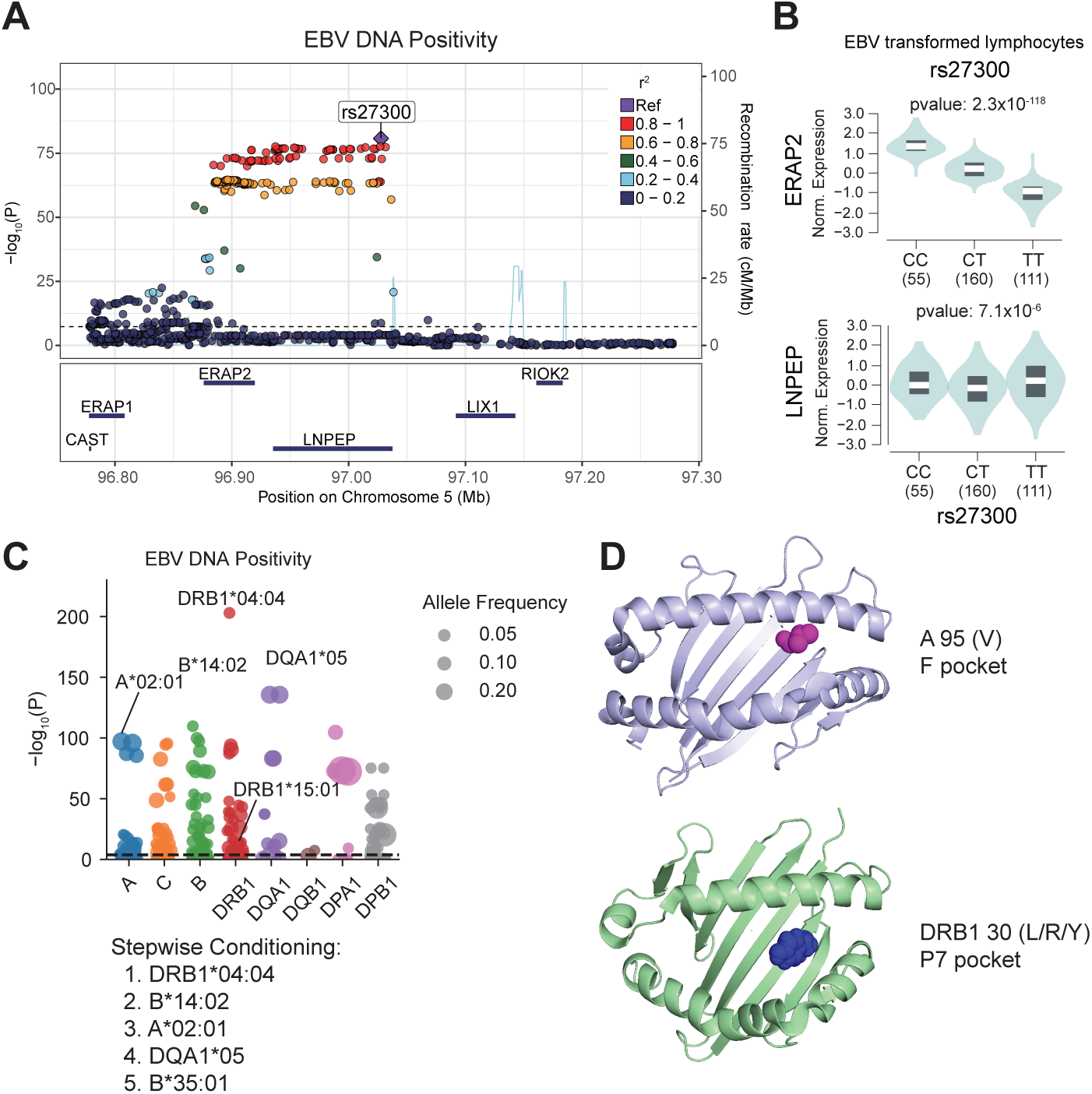
Association of HLA alleles and amino acid variants with EBV DNA positivity and viral load. (A) Locus plot of rs27300 in EBV Positivity GWAS. (B) eQTL plot of rs27300 in *ERAP2* and *LNPEP* in EBV-transformed lymphocytes. The eQTL data was downloaded from GTEx Portal.^122^ (C) Associations between HLA alleles and EBV DNA positivity. The size of each point corresponds to the allele frequency, and the top associated alleles are labeled. The alleles highlighted by conditioning were also shown. HLA association analysis using All of Us European participants was performed by imputing classical HLA alleles with SNP2HLA. (D) Amino acid residue associated with EBV DNA positivity/viral load in HLA-A*02:01 (top) and HLA-DRB1*04:04 (bottom). The structures were downloaded from the PDB (3MRF for HLA-A*02:01 and 7nzf for HLA-DRB1*04:04).

We also investigated the associations with HLA alleles and amino acid usage and EBV susceptibility. Using All of Us participants of European ancestry, classical HLA alleles were imputed with SNP2HLA,^51^ and their associations with EBV activity (DNA positivity and DNA viral load) were examined. In both traits, *HLA-DRB1*\*04:04 showed the most significant association. After conditioning analysis, the top alleles associated with EBV DNA positivity were *HLA-DRB1*\*04:04, *B*\*14:02, *A*\*02:01, *DQA1*\*05, and *B*\*35:01, while those for EBV DNA load were *HLA-DRB1*\*04:04, *A*\*02:01, *B*\*35:01, *DQA1*\*05:01, and *B*\*14, showing a broadly consistent pattern between the two traits (Fig. 2C and S6B, 7, 8, Supplementary Tables 12, 13). While to a lesser degree, EBV DNA positivity was also seen with the strongest MHC associated allele, *DRB1*\*15:01. Furthermore, an amino acid–level association analysis identified multiple residues significantly correlated with EBV activity (Figs. 2C, S6B, S7, S8, and Supplementary Tables 14, 15). Notably, valine at position 95 of *HLA-A*\*02:01 (A95V), detected in both positivity and viral load analyses, was located within the F pocket of the peptide-binding groove, while position 30 of HLA-DRB1 (L/R/Y variants), detected in the viral load analysis, was located in the P7 pocket (Fig. 2D). Both residues are situated within key structural regions that determine peptide-binding specificity, suggesting that EBV-associated HLA signals may reflect functional variation in peptide-binding pockets influencing viral antigen presentation and immune recognition.

Finally, we performed a detailed characterization of ERAP2 function. ERAP2 preferentially trims peptides bearing R/K at the N-terminus, resulting in the exposure of downstream residues such as F/Y/W at the N-terminus. Consistent with this, re-analysis of immunopeptidome data^52^ under ERAP2 perturbation reproduced a similar N-terminal motif (Fig. S6C). These results indicate that the effect of ERAP2 may not be restricted to specific viral antigens but broadly shapes the peptide repertoire, while potentially contributing to the presentation of a subset of EBV-derived antigens. Taken together, these findings suggested that antigen presentation is a key component of EBV regulation and a hotspot of susceptibility.

### Tissue-, Cell-, and Niche-Specific Heritability Analyses

To elucidate the biological contexts underlying host control of EBV activity, we performed partitioned heritability analysis using stratified LD score regression (S-LDSC),^53^ which infers tissue-level enrichment based on external functional annotations. Both EBV positivity and EBV DNA load exhibited marked enrichment in immune-related tissues (Fig. S9A). For EBV positivity, significant heritability enrichment was observed in spleen, peripheral blood, lymph nodes, synovial fluid, and lung, suggesting that systemic and mucosal immune environments contribute to host susceptibility to EBV activity. In contrast, EBV DNA load showed its strongest enrichment in EBV-transformed lymphocytes, followed by spleen, lymphoid tissue, and blood, indicating a link between viral load regulation and B cell transformation processes.

We next sought to interpret the polygenic signals identified in the EBV GWAS in the context of specific cell types and spatial niches using integrated single-cell and spatial transcriptomic analyses. We first applied single-cell disease relevance score (scDRS)^54^ to single-cell RNA-seq data from PBMCs of EBV-positive and control individuals,^55^ analyzing all cell types jointly. This analysis revealed enrichment of EBV-associated polygenic signals in cytotoxic lymphocyte populations, including CD8 effector memory T cells, NK cells, and γδ T cells (Fig. S9B).

Using the same dataset, we then focused the analysis on B-cell lineages that are directly involved in EBV infection. The dataset included publicly available single-cell RNA-seq profiles from patients with infectious mononucleosis and hemophagocytic lymphohistiocytosis (HLH). Existing pipelines based on Cell Ranger remove overlapping reads, resulting in substantial loss of reads from the compact EBV genome, and they may misalign reads with sequence similarity to human transcripts.^56^ To overcome these limitations, we developed VIRTUS3, an analysis framework optimized for viral detection in single-cell data (https://github.com/yyoshiaki/VIRTUS3). In this workflow, reads are first aligned to the human genome using Cell Ranger. Unmapped reads are then recovered and re-aligned to viral genomes using Alevin. This pipeline can filter out reads with stringent similarity to the human genome, and manipulate the reads aligned to multiple genes on viral genomes. This approach enabled the detection of 1.84-fold more EBV-positive cells compared with conventional methods (Figs. 3A-C). EBV-positive cells were predominantly detected among plasmablasts, followed by B memory and B intermediate cells, whereas naïve B cells showed almost no EBV positivity (Fig. 3D). Using scDRS, we further evaluated the cell-type–specific distribution of EBV DNA-associated polygenic scores. In memory B cells, polygenic scores were significantly elevated in EBV-positive compared with EBV-negative cells (*p*=3.0×10^-5^), while no difference was observed in plasmablasts (Figs. 3E and S9C). These findings indicate that host genetic control of EBV activity is exerted primarily within the memory B-cell compartment rather than at the terminally differentiated plasmablast stage.

**Figure 3.**
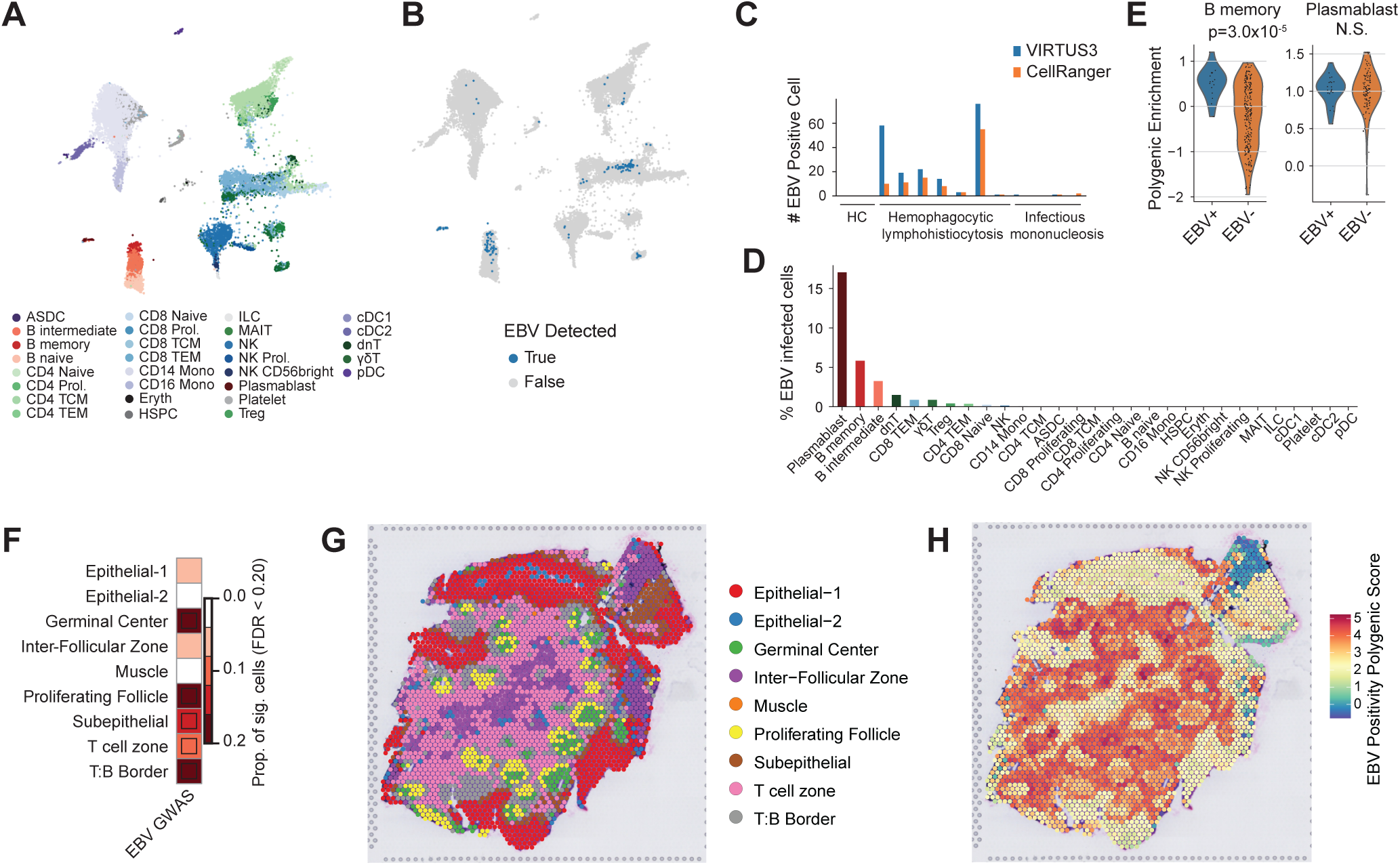
Tissue-and cell-type–specific heritability enrichment for EBV positivity and viral load. (A) UMAP visualization of PBMC single-cell RNA-seq data from EBV-positive and control individuals (DRA017005),^55^ showing major immune cell clusters. (B) Cells with detected EBV transcripts identified by VIRTUS3 (blue) overlaid on the UMAP plot. (C) Comparison of the number of EBV-positive cells detected by VIRTUS3 versus Cell Ranger across samples. (D) Distribution of predicted cell types among EBV-positive cells, highlighting enrichment in plasmablasts and memory B cells. (E) Polygenic scores derived from the EBV DNA positivity GWAS in B memory and plasmablast populations. The Mann-Whitney U test was applied. (F) Region-level enrichment of EBV DNA positivity polygenic signals in tonsils^117^ estimated by scDRS-spatial. Heatmap colors depict the proportion of significant cells (*FDR*<0.2) evaluated using scDRS^54^ Squares denote significant disease associations (*FDR*<0.05). (g and h) Spatial transcriptomic map of human tonsil tissue showing annotated microanatomical regions and EBV DNA positivity polygenic scores across the same tissue section.

We next investigated the immune microenvironments in which host genetic factors regulating EBV activity are concentrated by applying scDRS-spatial, a spatial heritability framework we previously developed^57^. At the tissue level, polygenic signals showed the strongest enrichment in lymph nodes and tonsils, confirming that these lymphoid tissues represent major sites of EBV regulation (Fig. S9D). We then analyzed the spatial localization of genetic signals within tonsillar tissue, a key reservoir for EBV infection, latency, and reactivation. Polygenic signals for both EBV DNA positivity and viral load were highly enriched within the germinal center, T:B border, and proliferating follicle regions, which are immune niches characterized by active B cell responses and dynamic B–T cell interactions (Figs. 3F–H and S9E). These findings indicate that host genetic factors controlling EBV activity act predominantly within lymphoid microenvironments supporting T cell–dependent B cell activation. In particular, the germinal center and T:B border emerge as key immunological niches where genetic variation may influence EBV persistence and immune control.

### EBV DNA detection in blood from MS patients

We next examined whether EBV DNA levels are directly associated with MS by qPCR. We compared whole-blood EBV DNA positivity between treatment-naive MS patients and non-MS individuals collected at the Yale MS Center. EBV DNA was detected in 11.5 percent of MS patients (27/234) and 5.26 percent of controls (9/171), showing a higher proportion in MS (Fig 4A; OR=2.35, *p*=0.0033, tested by two-sided Fisher’s Exact Test). To more rigorously assess this association, we conducted a meta-analysis of 11 studies encompassing 1,740 individuals and 525 EBV DNA–positive events (Figs. 4A, S10, and Table S16. The fixed-effect model yielded an odds ratio of 1.85 (95% *CI*=1.48–2.33, *p*<0.0001), and the random-effects model produced a nearly identical estimate (*OR*=1.85, 95 % *CI*=1.39–2.45, *p*=0.0007). Heterogeneity was low (*I²* = 11.0 percent), indicating consistent effects across studies. Although the effect size in MS is more modest than in other autoimmune diseases such as SLE or RA, where odds ratios typically range from 3.5 to 5,^27,58^ well-controlled case–control comparisons consistently demonstrate higher circulating EBV DNA in MS. This pattern reinforces the concept that EBV activity is modestly elevated around the time of clinical MS onset, but critically, it is important to note that this does not give insight into EBV activity at the time of initial infection when the virus may trigger disease initiation many years before clinical manifestation of CNS lesions.

**Figure 4.**
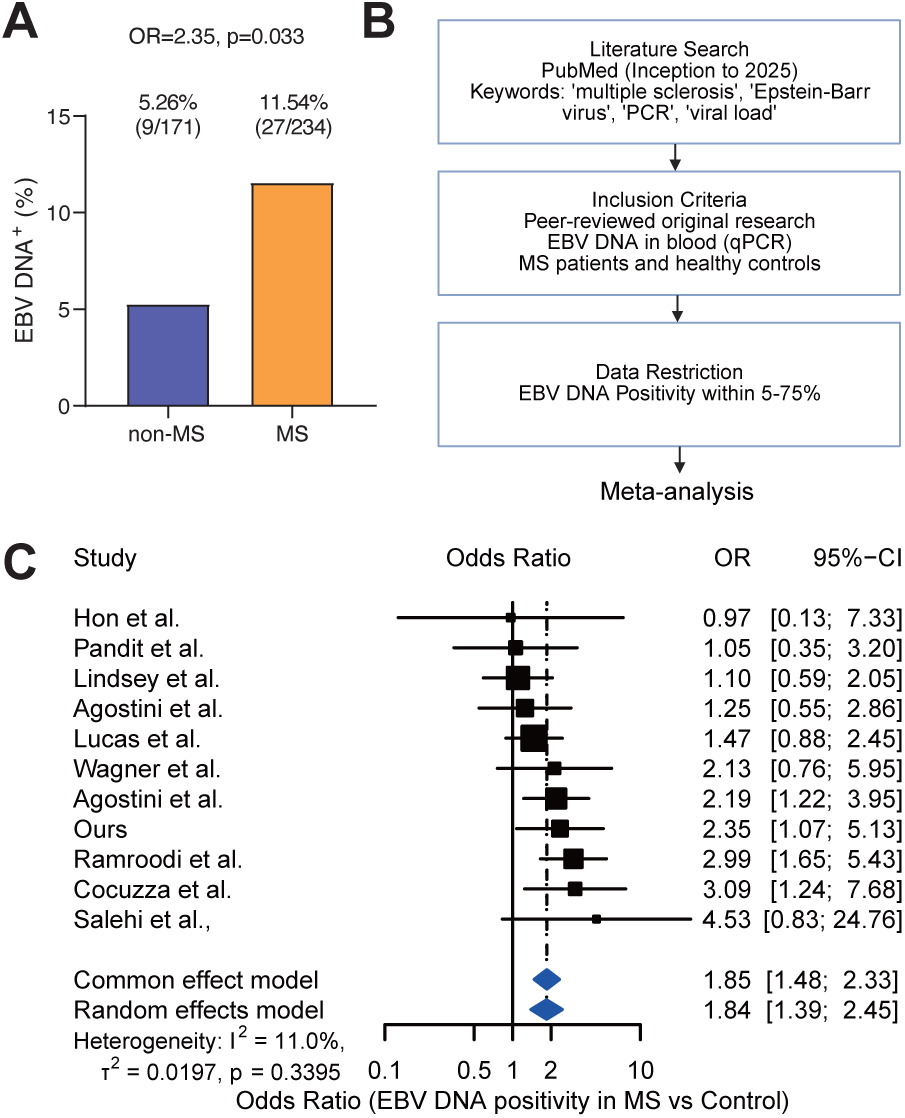
EBV DNA positivity is more frequent in patients with MS (A) Frequency of EBV DNA+ in MS and non MS individuals in Yale cohort. (B) Scheimatic view of MS EBV DNA meta-analysis. (C) Forest plot showing OR and 95 percent confidence intervals for EBV DNA positivity comparing patients with multiple sclerosis (MS) and non-MS controls across published studies and our cohort. Squares indicate study-specific ORs with sizes proportional to study weights, and horizontal lines represent 95 percent CIs. The diamonds indicate the pooled effect estimates under fixed-effect and random-effects models. Quantification of heterogeneity: τ*^2^*=0.0169, τ=0.1301, *I^2^*=9.8 percent [0.0 percent; 49.4 percent], *H*=1.05 [1.00; 1.41]. A funnel plot is shown in Fig. S12.

### Single-cell resolution cellular dynamics in EBV-infected B cells in people with MS

We next sought to directly characterize the interaction between EBV infection and MS-related genetic programs by performing single-cell analyses of PBMCs from MS and healthy individuals with EBV detection at single-cell resolution. A comprehensive survey of public scRNA-seq datasets revealed virtually no EBV-positive cells.^59^ This was unlikely to reflect a true biological absence but rather a technical limitation of conventional single-cell RNA-seq, which captures only polyadenylated transcripts and therefore fails to detect the major latent EBV noncoding RNA *EBER1*, which lacks a polyA tail. To overcome this limitation, we introduced a spike-in probe enabling specific capture of *EBER1*, which is a similar approach to EBV-seq.^60^ Yet even with this improved method, EBV-infected cells remained exceedingly rare, with none detected in PBMCs (0 of 51,950 cells from 3 donors) and only three cells detected in tonsils (3 of 134,073 cells from 12 donors) from both healthy donors and MS patients, underscoring a fundamental limitation of single-cell approaches when the infected population is extremely sparse (Fig. S13).

Previous studies have shown that blood EBV DNA load strongly correlates with the number of *EBER1*-positive cells,^61^ suggesting that EBV DNA–positive samples may enable the recovery of EBV-infected cells at single-cell resolution. We therefore analyzed banked frozen PBMCs from EBV–positive individuals screened by qPCR analysis. We again performed single-cell RNAseq with the *EBER1* spike-in probe, prioritizing EBV DNA–positive MS and non-MS samples (Fig. 5A). With this technique, we could detect 1,069 DNA–positive cells from 470,989 B cells (0.23%) (Figs. 5B and C, Supplementary Table 17). As expected, the number of EBV–positive cells were positively correlated with qPCR-based EBV DNA load (*rho*=0.55, *p*=5.0×10^-4^, Fig. 5D). The EBV–positive cells were predominantly memory B cells, antibody-secreting B cells (AbSCs), naive B population expressing Type 1 interferon signature genes (Bnaive MX1), and ASC precursor naive B cells (Figs. 5E and S14B). Notably, most of the EBV–positive cells expressed latent EBV genes, including *EBER1, LMP-1, and LMP-2A*, while some antibody-secreting B cells expressed lytic genes even in high EBV DNA load individuals (Figs. 5E and S14B). Interindividual variation was observed in the distribution of EBV-positive cells (Fig S14B). For example, some individuals showed enrichment in switched memory B cells (Bsm cells) and ASCs, others in T-bet^+^ CD11c^+^ Atypical B cells (atBCs), and others in naive B cells and ASCs, indicating several distinct distribution patterns. In most EBV–positive cells, the transcriptional program was dominated by latent EBV genes. However, in a subset of individuals, cell populations expressing *BARF0* and *A73* were observed, and a few individuals also harbored rare cells expressing early lytic genes.

**Figure 5.**
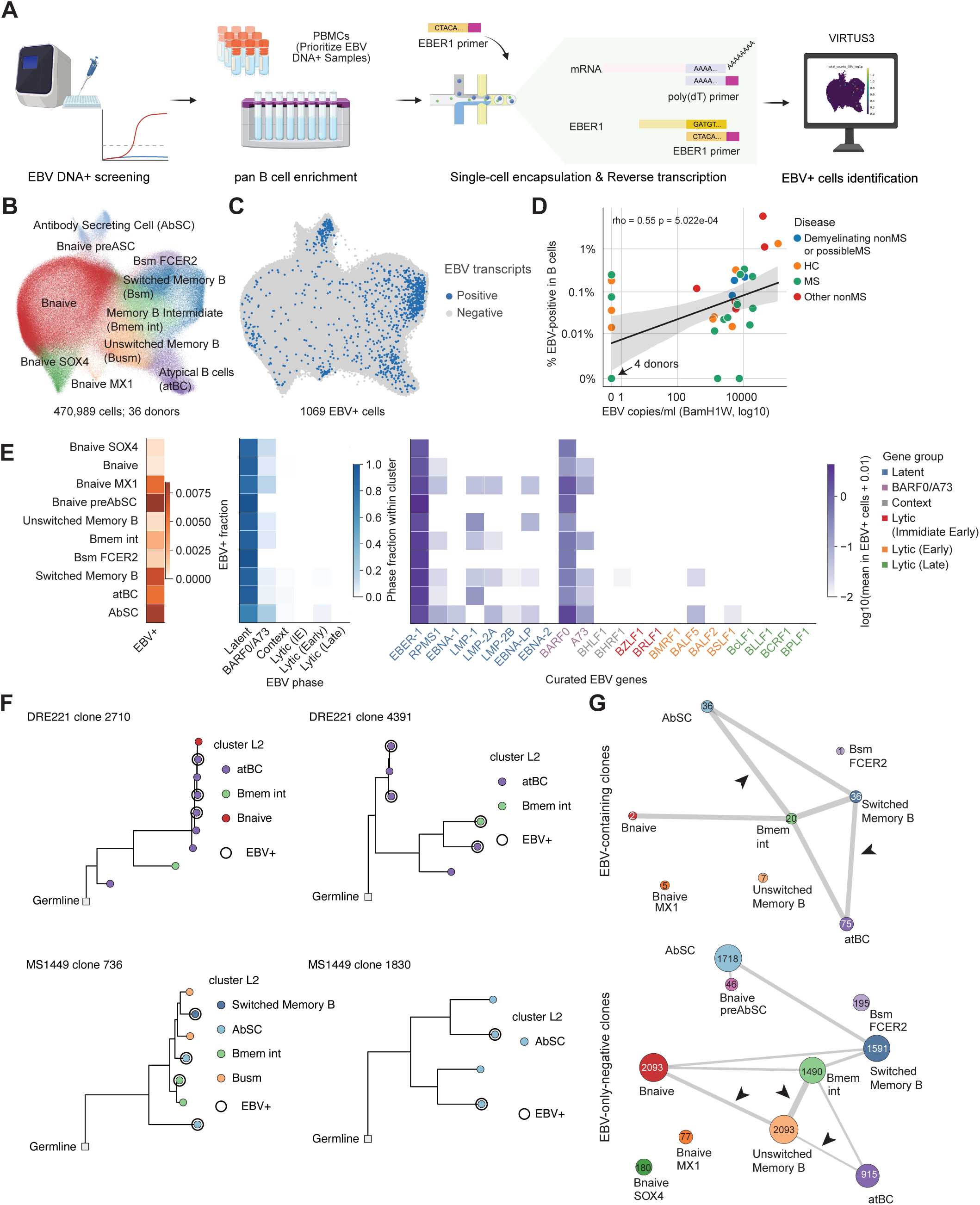
Single-cell EBV detection from people with MS and healthy individuals (A) Schematic view of scRNAseq experiments. (B and C) UMAP embedding of B-cell subsets from PBMCs (B) and EBV–positive cells (C). (D) Relationship between EBV copies per mL (qPCR) and the percentage of EBV⁺ B cells, stratified by disease status (MS vs non-MS). The shaded region denotes a 95 percent confidence interval. Spearman’s *rho* and *p-value* are shown in the figure. (E) Heatmap of viral transcript detection across B-cell subsets, displaying the number of cells expressing canonical EBV latent and lytic transcripts and aggregated EBV gene expression. (F) Representative clonal trees including EBV–positive cells. (G) Clonal network representations of clones containing EBV–positive cells and clones lacking EBV–positive cells. Node size indicates the number of cells in each cluster, and edge width indicates inter-cluster BCR repertoire similarity, defined as 1 - Jensen-Shannon divergence. Only the top 20% of edges by similarity are shown among clones with n >= 2 cells. Representative differential edges are marked by arrow heads.

EBV also affected BCR characteristics (Figs. S14B-D). Somatic hypermutation was increased in EBV–positive cells in Bnaive MX1 and Bmem intermediate (Bmem int). *IGHG3* and *IGHG1* were more frequent, and *IGHM*, *IGHD, and IGHA2* were less frequent. EBV–positive cells tended to be more expanded, especially in Bnaive MX1, Bmem int, atBCs, and AbSCs. Clonal analysis identified multiple clones derived from the germline BCR sequence containing EBV-positive cells (Fig. 5F and G). These clones showed diverse cellular compositions, including clones enriched for atBCs and Bmem int, clones composed almost entirely of AbSCs, and clones consisting of both AbSCs and memory B cells. Integrative analysis showed the differentiation pattern observed in conventional EBV-negative clones, in which intermediate B cells progressed through unswitched memory B cells toward memory B cells, was not evident in EBV-positive clones. In contrast, shared clonal relationships linking atBCs to switched memory B cells were relatively prominent among EBV-positive clones. These findings suggest that, although the distribution of EBV-infected cells varied across individuals, EBV transcription in peripheral blood was dominated by latency-associated genes, and EBV infection may influence B-cell states and differentiation trajectories.

Next, we examined how EBV-positive B cells alter their transcriptional programs. Pathway analysis revealed marked upregulation of BCR signaling, NF-κB signaling, antigen processing and cross presentation in memory B cells and viral mRNA translation in antibody-secreting cells (Fig. 6A, Supplementary Table 19). Importantly, focused analysis of cytokines and receptors involved in immune interactions (Fig. 6B) showed increased expression of *IL12A* and *IL23A* in EBV-infected memory B cells, which promote Th1 and Th17 differentiation, together with reduced expression of the immunoregulatory cytokine *IL10* and its receptor *IL10RA*. EBV-positive cells also upregulated *IL16*, *IL6ST* (gp130), and *IL6R*, molecules implicated in MS pathogenesis. While *CXCR4* was downregulated, molecules involved in recruiting and activating pathogenic T-cell subsets, including *CCR6* (Th17-associated) and *CXCR3* (Th1-associated), were also increased. In addition, EBV-positive cells showed elevated expression of receptors crucial for B-cell differentiation and survival, including *CXCR5*, *TNFRSF13B* (TACI), *TNFRSF13C* (BAFF-R), *TNFRSF14* (HVEM), and *TNFRSF17* (BCMA), several of which have prior genetic links to MS. Finally, Bmem int and Bsm upregulated T-cell help molecules, including *CD40*, *CD86*, and *HLA class I and class II*. Together, these findings suggest that EBV-infected memory B cells may contribute to immune tolerance breaking by promoting Th1/Th17 polarization while weakening regulatory immune circuits.

**Figure 6.**
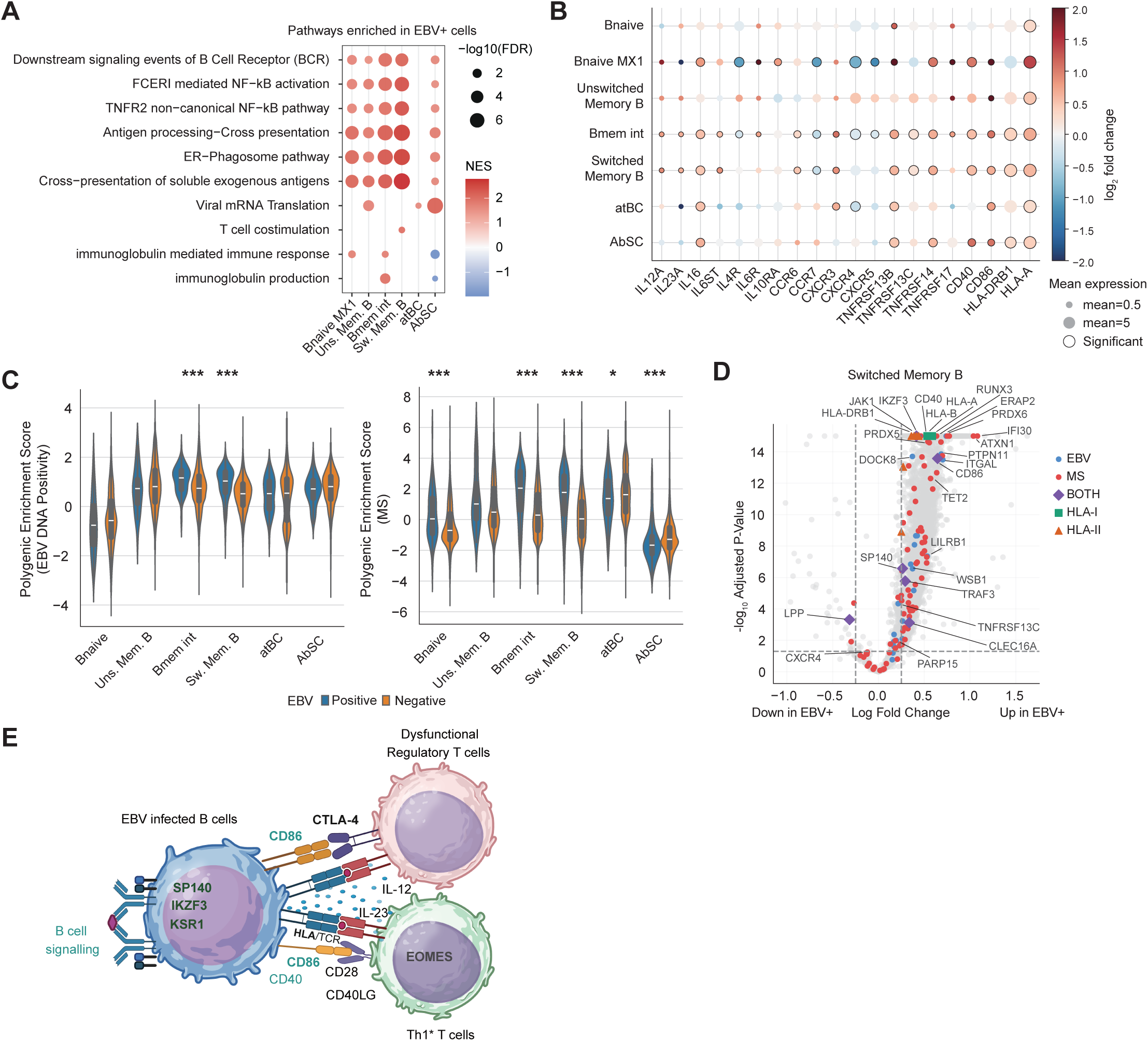
Gene program changes in EBV-positive cells (A) Gene Ontology (GO) enrichment analysis comparing EBV–positive versus EBV–negative cells for each B cell population. Dot size represents gene ratio, and color indicates adjusted *p*-value. (B) Dot plot showing expression level and log_2_ fold change of cytokines, chemokine receptors, and immunomodulatory molecules differentially expressed in EBV–positive cells. Differentially expressed genes are highlighted in bold circles. (C) scDRS-based polygenic score enrichment for EBV DNA positivity (left) and MS onset genes (right) across B-cell subsets, comparing EBV–positive and EBV–negative cells. A statistical test was performed by the Mann-Whitney U test. *: p_adj_ < 0.05, **: p_adj_ < 0.01, ***: p_adj_ < 0.001. (D) Volcano plot of differentially expressed genes in EBV–positive versus EBV–negative switched memory B cells. Genes highlighted in the EBV and MS GWAS are annotated. (E) Schematic view of genetic EBV susceptibility in EBV-infected memory B cells and suggested T cell involvement. Genes in bold font are genes highlighted in EBV DNA GWAS.

Finally, we investigated how host genetic factors interact with EBV infection across B-cell subsets. Using scDRS to evaluate EBV GWAS signals, we found that the EBV DNA positivity–associated polygenic signal was significantly enriched in EBV-positive B naïve and switched memory B cells, whereas no meaningful enrichment was observed in antibody-secreting cells (Fig. 6C). These findings suggest that early infection responses in naïve B cells and persistent infection in switched memory B cells represent key cellular contexts in which EBV regulation is exerted. We then performed a similar analysis using MS onset–associated gene sets. Polygenic scores for MS were markedly elevated in EBV-positive unswitched memory B cells and switched memory B cells, while antibody-secreting cells showed a reduction in MS-related signal (Fig. 6C). Specifically, MS risk gene expression was increased within EBV-infected cells in Bnaive, atBCs, and memory B cells, indicating that EBV-infected B cells constitute a biological context where MS risk genes are preferentially engaged. Notably, many MS susceptibility genes, EBV susceptibility genes, and shared loci between the two GWAS, including *HLA class I and II* molecules, *IKZF3*, *SP140*, *CD86*, *TRAF3, and CLEC16A*, showed increased expression in EBV-positive switched memory B cells (Fig. 6D). These observations indicate that EBV infection activates a gene network that intersects with MS susceptibility specifically within defined B-cell subsets. Taken together, these results demonstrate that EBV infection enhances both viral polygenic signals and MS susceptibility gene expression in select B-cell subsets, revealing a cellular setting in which viral regulation and autoimmune genetic risk converge.

## Discussion

Here, we developed a pipeline to quantify EBV DNA from whole-genome sequencing data and applied it to population-scale cohorts. A cross-ancestry GWAS of EBV DNA positivity in 617,186 individuals identified 39 susceptibility risk loci. We validated this finding in our independent cohort and found that qPCR-confirmed EBV DNA-positive individuals were markedly enriched in the top decile of EBV polygenic risk scores containing these newly discovered loci. Intriguingly, there was a significant overlap with genetic variants associated with MS, RA, and SLE susceptibility alleles, and Mendelian randomization analyses further supported a causal influence of EBV activity on MS, largely driven by HLA variation. By establishing a single-cell RNA-seq method optimized for *EBER1* detection, we found that EBV RNA was detected primarily in memory, atypical, and antibody-secreting B cells, with interindividual heterogeneity. Notably, EBV was predominantly in the latent phase even in high EBV DNA load individuals. BCR lineage analysis revealed that memory B cell differentiation is altered by infection. EBV-infected memory B cells, including switched memory B cells strongly upregulated cytokines and risk genes from both the EBV and MS GWAS. These data demonstrate that EBV–infected B cells are a key modulator of T cells and that MS susceptibility genes are preferentially activated within EBV-infected B cells (Fig. 6E). This identifies a genetic and immunologic mechanism by which EBV infection leads to pathologic activation of B cells in a genetically susceptible individual, driving the multitude of immune dysfunctions observed in MS with subsequent initiation of disease.

MS is the autoimmune disease for which EBV is most strongly implicated as a necessary environmental factor, and both tissue and cell specificity of EBV regulation further align with MS-specific immunobiology in which memory B cells drive CNS infiltration.^62^ Moreover, it has been clearly demonstrated that the EBV virus harbors sequences that are cross-reactive with myelin epitopes, including GlialCAM and anoctamin-2, that are likely also playing a critical role in the pathogenesis of MS^18–20^. Thus, EBV uniquely may drive activation of myelin-reactive T cells both by breaking immune tolerance and by presenting cross-reactive antigens.

Although EBV DNA showed no positive association with MS in our PheWAS,^63^ a well-controlled treatment-naive MS cohort and the case-control meta-analyses showed elevated EBV DNA levels in MS patients. This discrepancy is likely explained by the time course of EBV involvement in the onset of MS. Specifically, EBV seroconversion occurs an average of 7.5 years before clinical onset,^12^ MS diagnosis requires CNS lesions disseminated in time and space,^64^ and a prodromal period precedes diagnosis.^65^ As a result, EBV load at the time of diagnosis is likely to be partially deflated. Furthermore, individuals with MS in cohorts such as All of Us include patients receiving long-term immunotherapy and those many years after diagnosis, making it difficult to capture EBV activity during the true initiation phase. These considerations highlight the importance of examining newly diagnosed, treatment-naïve patients or MS high-risk individuals prior to disease onset to accurately evaluate EBV activity in early MS.

Recent population-scale studies established the feasibility of quantifying EBV DNA from WGS data and highlighted genetic factors for controlling EBV persistence.^35,36^ However, the relationship between EBV activity and MS appears to depend on the level of genetic resolution being examined. At the broad polygenic level, the association with MS was modest, consistent with our own observations that diffuse genome-wide correlation is limited.^36^ By contrast, when focusing on lead EBV-associated loci, we observed clear overlap with established MS susceptibility loci. One plausible explanation is that the relationship between EBV control and MS is not captured by a uniform genome-wide effect, but rather by a more selective set of shared loci with heterogeneous directions of effect. We therefore view prior studies and our results as complementary: earlier work defined the general host-genetic architecture of EBV persistence, whereas our study identifies a more specific overlap at highlighted loci and further localizes that convergence to EBV-infected B-cell states using single-cell and spatial transcriptomic analyses.

Our large-scale WGS analysis also clarified the broader epidemiology and biology of EBV infection across populations. In the United States, individuals of African, Latino/admixed American, and Asian ancestry showed higher EBV positivity rates, consistent with serology-based findings.^66^ Female individuals showed lower EBV DNA positivity despite a higher prevalence of autoimmunity. This pattern suggests that sex differences in EBV susceptibility and the host immune response to EBV may shape autoimmune risk. We also found that a small subset of “super-carriers” accounted for a disproportionate fraction of EBV DNA detected in the population.

Cross-ancestry GWAS clarified the host genetic architecture underlying EBV activity. Strong enrichment of associations in the HLA region, in T-cell–related pathways including association in *CTLA4*, *EOMES*, and *CD86*, and at the T–B border of secondary lymphoid tissues, underscores the central role of B cells in antigen presentation to T cells in EBV regulation. EBV employs multiple immune-evasion strategies, including BNLF2a-mediated inhibition of TAP1.^67^ Here, we showed that HLA polymorphisms shape antigen presentation efficiency and directly influence EBV activity. *HLA-DRB1*\*04:04, a known risk allele for RA and MS,^68,69^ and an allele that presents myelin-specific TCRs in MS patients,^70^ was associated with higher EBV activity. The risk allele *HLA-DRB1*\*15:01 was also associated with higher EBV antigen presentation, though to a lesser extent than the *HLA-DRB1*\*04:04 allele, suggesting other factors contribute to the genetic risk associated with these MHC haplotypes. In contrast, *HLA-A*\*02:01 was associated with reduced EBV activity and is protective against MS^71^ and EBV⁺ Hodgkin lymphoma.^72^ HLA molecules shape antigen-presentation preferences and thymic TCR repertoire formation.^73^ In addition, components of the antigen-processing pathway, including *ERAP2*, emerged as important regulators of EBV control. The reanalysis of immunopeptidomic data indicates that *ERAP2* is a key regulator of peptide presentation via MHC class I and EBV clearance. *ERAP2* allotype is also associated with MS susceptibility.^74^ Our findings indicate that HLA diversity and related genes exert strong effects on EBV susceptibility and that several EBV-associated alleles overlap with autoimmune risk alleles, suggesting shared genetic pathways linking EBV control and autoimmunity.^75^ Moreover, PRS and MR analysis highlighted the HLA contribution to autoimmune diseases, including MS.

Non-HLA loci further underscore that EBV regulation relies heavily on intrinsic programs of the host B cell. Genes in which the direction of effect was concordant between EBV DNA positivity and autoimmune disease risk, including *RSBN1*, *SLAMF7*, *CTLA4*, *EOMES*, *CD86*, *TP63*, *TRAF3*, *CLEC16A*, and *IKZF3*, converged on pathways governing B-cell differentiation, germinal-center dynamics,^76^ NF-κB signaling,^77^ T cell effector function, and antigen presentation. This pattern suggests that B-cell differentiation and activation programs provide a shared biological foundation that simultaneously promotes EBV persistence and increases autoimmune susceptibility. In contrast, genes showing discordant effects, such as *SP140*, *SH2B3* and *PTPN11*, map to pathways involved in immunoregulatory thresholds and negative feedback, raising the possibility that trade-offs between antiviral responsiveness and autoimmune risk underlie their opposing directions of effect. Representative loci also suggested plausible mechanisms of EBV control: SP140 and IFI16 are linked to viral genome surveillance and restriction of viral gene activation,^78–80^ whereas TERT suggests a role for telomere biology in EBV replication and long-term survival of infected B cells. Together, these loci highlight a second layer of EBV control, intrinsic B-cell programs that operate in parallel with HLA-mediated antigen presentation.

Building on these findings, we further leveraged three key advances: improved single-cell detection of viral transcripts, an optimized bioinformatics pipeline, and sample prioritization based on EBV DNA by qPCR, to achieve the direct identification of EBV-infected cells in patients with MS at single-cell resolution. By analyzing a large cohort enriched for untreated, newly diagnosed MS cases, we were able to capture EBV-positive cells during the clinically relevant early phase of disease. These infected cells were concentrated within B switch memory cells, atBCs, and ASCs. The predominance of latency-associated EBV transcripts suggests that latent-phase viral programs may be particularly relevant to MS-related immune dysregulation. Indeed, LMP1 and LMP2A are known to mimic or rewire CD40 and B-cell receptor signaling, respectively, and EBNA2 binds MS susceptibility loci in a genotype-dependent manner.^81^ This is consistent with the observation that EBV-positive cells showed altered B-cell differentiation trajectories. Atypical B cells have recently emerged as an important component of MS immunopathology,^82,83^ although their persistence and developmental fate remain context-dependent.^84^ In our data, shared clonal relationships between atBCs and B switched memory cells were relatively prominent among EBV-positive clones, suggesting that EBV infection may support the persistence of an atBC-associated program or facilitate its transition into longer-lived switched memory states.^85^

EBV-infected B cells showed a coordinated activation of MS genetically mediated immune communication pathways, including upregulation of *IL12A* and *IL23A*, which drive Th1 and Th17 differentiation. IL-12 has been shown to diminish Treg suppressive capacity in inflammatory settings, shifting the balance toward effector responses.^86^ This pro-inflammatory shift was further reinforced by reduced expression of *IL10* and its receptor *IL10R*, consistent with impaired regulatory B-cell function described in MS.^87^ EBV-positive cells also increased *IL16*, *IL6ST* (gp130), and IL6R, molecules implicated in MS pathogenesis, and upregulated the chemokine receptors *CCR6* and *CXCR3*, which position B cells within Th17-and Th1-rich inflammatory niches rather than homeostatic environments. In parallel, EBV-positive B cells showed elevated expression of receptors crucial for B-cell differentiation and survival, including *CXCR5*, *TNFRSF13B* (TACI), *TNFRSF13C* (BAFF-R), *TNFRSF14* (HVEM), and *TNFRSF17* (BCMA). The upregulation of *CD86*, *CD40*, and *HLA class I/II* also supports the enhanced T cell help. Together, these findings indicate that EBV infection rewires B-cell immunobiology that is enhanced by genetic variants associated with risk of MS by enhancing T-cell help, promoting positioning within inflammatory niches, and reinforcing B-cell differentiation and survival programs relevant to MS.

Notably, both MS susceptibility genes and EBV activity–associated genes were highly expressed within these EBV-infected memory B cells. This represents the first direct evidence that host genetic risk for MS and genetic determinants of EBV activity converge within EBV-infected memory B cells. Given that memory B cells serve as potent, antigen-specific professional antigen-presenting cells, targeting this compartment provides a compelling biological rationale for the remarkable efficacy of B-cell–depleting therapies in MS.

Epidemiologic studies indicate that there is an approximately seven-year lag between EBV infection and the clinical onset of MS, and before there are clinical symptoms, patients are experiencing new, intermittent MRI lesions. This raises the question as to whether EBV is the initial trigger of disease, or viral reactivation leads to disease flare-ups later in the disease, or both. We hypothesize a mechanism by which EBV infection in genetically susceptible adolescents leads to a highly intense, acute activation of myelin-reactive T cells where overexpression of key immune pathways or “danger signals” in B cells, again mediated by the genetic architecture as demonstrated here, breaks immune tolerance in a stochastic manner, leading to a cascade of events initiating autoimmune MS.

Taken together, our work identifies the epidemiology of EBV across ancestries and elucidates the genetic basis of EBV B cell expression. By integrating multiple omics data, we mapped the key immunological “hotspots” where this regulation occurs. These findings support the existence of shared biological pathways linking EBV to autoimmune diseases, including MS. Notably, EBV-infected memory B cells and atypical B cells upregulated cytokines and costimulatory signals that influence T cell activation, IFNγ secreting Tregs, and regulators of B cell differentiation and survival, enhanced by MS susceptibility genes. EBV-infected memory B cells upregulated risk genes from both the EBV and MS GWAS, suggesting that EBV-infected B cells constitute a critical hub that modulates T cell responses while simultaneously activating MS susceptibility pathways within the B cell compartment. Together, these findings define a definitive genetic and cellular framework elucidating the potential molecular mechanisms causing MS.

## Data and code availability

GWAS summary statistics will be available at the GWAS Catalog and PGS Catalog upon acceptance. Locusplot for all lead SNPs and the adjacent region is available at 10.6084/m9.figshare.30712496. Single-cell RNAseq data generated in this study will be deposited in GEO and CZ CELLxGENE upon acceptance. Code generated for this work will be publicly accessible on GitHub upon acceptance. The pipeline used for EBV DNA detection from WGS is available at https://github.com/shohei-kojima/WGS_EBV_search. VIRTUS3 is available at https://github.com/yyoshiaki/VIRTUS3. All codes used for the analysis will be available on github (https://github.com/yyoshiaki/EBV_MS_2025_Manuscript) upon acceptance.

## Author contributions

Conceptualization, Y.Y., S.K., K.I., C.R., and D.A.H; data analysis, Y.Y., C.R., S.K., N.K., and H.L.C.; experiment, N.K. and J.M.; data curation and sample collection, L.Q., S.H., A.S., N.B.P, A.M. and E.E.L.; writing, Y.Y., N.K., C.R., S.K., E.E.L., G.J.F. and D.A.H.; supervision, Y.Y., T.S.S., K.I., E.E.L., G.J.F., and D.A.H.; funding acquisition, Y.Y., D.A.H., and G.J.F.

## Acknowledgement

This work was supported by NIH grants (P01 AI073748, U19 AI089992 U24 AI11867, R01 AI22220, UM 1HG009390, P01 AI039671, P50 CA121974, and R01 CA227473); the Race to Erase MS (to DAH); the National MS Society (to DAH and EEL); Department of Defense (HT9425-24-1-0522 to EEL); the Common Mechanisms in Autoimmunity Insight Award jointly funded by Breakthrough T1D, the Lupus Research Alliance, and the National MS Society (to YY), and Robert E. Leet and Clara Guthrie Patterson Trust Mentored Research Award, Bank of America, Private Bank, Trustee (to YY). YY is supported by JSPS Overseas Research Fellowships. NK is supported by the Sejong Science Fellowship (Overseas) from the National Research Foundation of Korea (RS-2024-00342077). CR is supported by an AAN/AHA Ralph L. Sacco Scholars Fellowship (AHA.24RSSPOST1328228), a Pilot Grant from the Claude D. Pepper Older Americans Independence Center at Yale School of Medicine (P30AG021342), a Pilot Award from the Yale Center for Brain and Mind Health, and a generous gift from the Paul and Sandra Montrone from the Penates Foundation. The illustrations were generated using BioRender.com.

## Methods

### Human Subjects

In this study, we analyzed whole-genome sequencing (WGS) data and linked electronic health record information from participants enrolled in the All of Us Research Program and the UK Biobank. All analyses were conducted in accordance with each data resource’s governance framework (All of Us: Registered Tier; UK Biobank: Application 58743), and only authorized researchers accessed the data within secure computing environments. For All of Us, WGS data derived from both blood and saliva were used, whereas UK Biobank provided WGS data derived from blood DNA. In both cohorts, all datasets were de-identified, and no personally identifiable information was available to researchers. Clinical information included ICD-9/10 diagnosis codes, age, sex, self-reported ancestry, and laboratory measurements, limited to variables approved for research use within each biobank.

A cohort of individuals at increased risk for MS was recruited at Yale University. This cohort study consists of (1) family members of MS patients and (2) asymptomatic individuals incidentally discovered to have morphologically suspicious lesions on brain MRI. Participants donate biospecimens and are prospectively followed. European-ancestry cohort members were selected for PRS analysis. A cohort of patients with MS used for qPCR and scRNAseq was also recruited at Yale School of Medicine.

### EBV DNA quantification from WGS

The human reference genome, GRCh38, which is used in AoU and UKB, contains the EBV decoy sequence, named chrEBV. To detect EBV reads in WGS samples, we investigated reads mapped to chrEBV stored in the CRAM files. A WGS sample can sometimes contain very abundant EBV reads due to viremia and have the potential to cause a heavy computational burden. To decrease the computational cost, samples with more than 350,000 reads mapped to chrEBV were randomly downsampled using samtools to 350,000 reads. Reads were then filtered to retain only proper pairs, passing standard quality control metrics, with duplicates, secondary, and supplementary alignments removed. Mapping length was calculated from the CIGAR string, and only reads with an aligned length of ≥145 bp were retained. The EBV genome carries a repetitive region, named BamHI W repeat (positions 12,000–35,355 bp). EBV reads derived from this region can be multi-mapping due to the repetitive nature of the BamHI W repeat region in the EBV reference genome. Our pipeline accounts for this repetitive nature and avoids counting such multi-mapping reads repeatedly. Finally, paired-end reads passing all filters were defined as EBV reads, and counts were summarized separately inside and outside the BamHI W repeat. In the case of analysis with downsampling, it estimates the number of EBV reads in the original CRAM file by accounting for the downsampling ratio.

### Association analysis of EBV DNA

We analyzed the association of EBV positivity with age_at_collection, genetically predicted ancestry, which is provided by All of US, and sex. Observations with missing values were excluded. Logistic regression (generalized linear model with binomial distribution and logit link) was fitted separately for blood samples and saliva samples. The model was specified as:

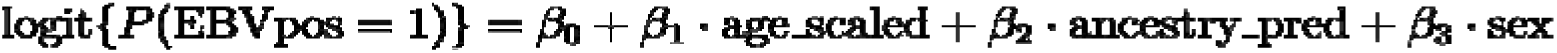

To facilitate interpretation, age was scaled in 10-year units (age_scaled = age_at_collection / 10). European ancestry (eur) and male sex were used as reference categories. Robust (HC3) standard errors were applied. Coefficients were reported as odds ratios (ORs) with 95% confidence intervals. Analyses were conducted in Python using the package statsmodels. Logistic regression models were estimated using statsmodels GLM (Binomial, logit) with robust HC3 standard errors.

### Generalized Pareto distribution (GPD) analysis

To assess the tail behavior of EBV DNA distributions, we applied a peaks-over-threshold (POT) approach using the Generalized Pareto distribution (GPD). Normalized EBV DNA reads (normalized by the number of human-genome coverage) were used. For each sample type (blood, saliva), the 95th percentile of the distribution was chosen as the threshold u, and exceedances above u were modeled with the GPD parameterized by shape (ξ) and scale (β). Parameter estimation was performed by maximum likelihood, and 95% confidence intervals of ξ were obtained via profile likelihood. For conceptual illustration, simulated GPD curves with ξ =-0.3, 0, 1.0, and a common scale were generated to highlight the difference between finite, exponential, and heavy-tailed distributions.

### Genotype Data Processing and Quality Control

Genotype data were processed independently for the All of Us Research Program^37^ and the UK Biobank,^38^ following harmonized quality control procedures to enable cross-cohort analyses. Unless otherwise noted, equivalent filters and thresholds were applied across both datasets. The variant set for downstream analyses was defined based on the All of Us ACAF PGEN callset and applied consistently to both cohorts.

For the analysis of All of Us, we utilized the All of Us ACAF PGEN callset, which includes variants with “AF>1% or AC>100 in any ancestry,” filtered using the FT flag. Participants were excluded if DNA was derived from saliva, retaining only blood-derived samples.

Following previous studies, we removed individuals with immunodeficiency disorders (ICD-10 codes D80–D89) or those with ‘ID_057.1’ (Human immunodeficiency virus infection) in PhecodeX. Related and flagged individuals were excluded using the All of Us auxiliary files (relatedness_flagged_samples.tsv and flagged_samples.tsv). We retained only samples with inferred sex chromosomes XX or XY, sex at birth recorded as male or female, and available age-at-collection information.

Variant-level quality control was performed per ancestry with the following parameters: MAF between 0.01 and 0.99, Hardy–Weinberg equilibrium (HWE) p>1×10^⁻^ (using keep-fewhet), biallelic variants only, and a genotype missingness rate <5%. An ancestry-specific variant list was generated for each group, and the union across all ancestries and cohorts was used for subsequent analyses.

For the UK Biobank cohort, EBV DNA was quantified from WGS data using the same pipeline as All of Us, counting reads mapping to the EBV reference genome (chrEBV). EBV read counts were normalized by mean sequencing coverage and log10-transformed (log10[normalized reads + 1]) for the quantitative trait analysis; a binary EBV DNA positivity variable was defined as having at least one EBV-mapped read. Sample-level quality control included: retention of individuals of European genetic ancestry based on the UK Biobank genetic ethnicity field (Data-Field 22006); exclusion of one member from each pair of related individuals (*kinship coefficient*≥0.0442) using the UK Biobank relatedness file; exclusion of individuals with sex chromosome aneuploidy or discordance between reported and genetic sex; removal of WGS preparation plates with unusually high EBV positivity rates (>28%); and exclusion of individuals in the top 1% of normalized EBV reads. As in All of Us, we excluded individuals with immunodeficiency disorders (ICD-10 codes D80–D89) or HIV infection (ICD-10 codes B20–B24), as well as those taking immunosuppressant medications at baseline (Data-Field 20003). Principal components were computed on unrelated European individuals and projected onto all European samples. For GWAS, we used imputed genotype data from UK Biobank. For REGENIE Step 1, we used directly genotyped variants after quality control (*MAF*≥0.05, *genotype missingness*<5%, *MAC*≥40, HWE *p*>1×10⁻²[, biallelic SNPs only) and LD pruning (window 200 kb, step 50 variants, *r^2^*<0.2), yielding 346,104 variants. For REGENIE Step 2 association testing, we applied filters of MAC ≥ 40 and imputation INFO ≥ 0.8. For the meta-analysis, we restricted to the same set of variants defined by the All of Us ACAF callset by matching on chromosome, position, and alleles.

### Genome-Wide Association Analysis and meta-analysis

Genome-wide association analyses were performed using REGENIE^40^ with comparable parameters across cohorts/population. In step 1, LD-pruned variants were selected using ancestry-specific thresholds.^88^ For individuals of African ancestry, we used --indep-pairwise 1000 100 0.15; for non-African ancestries, --indep-pairwise 1000 100 0.3. Covariates included the first 20 principal components, sex at birth, and age at collection (including quadratic and age×sex interaction terms), with additional cohort-specific covariates such as sequencing batch metrics, fasting time, lymphocyte percentage, smoking status, time of blood draw, and month of collection to account for technical and seasonal variation. Step 2 analyses were restricted to SNPs that passed QC in at least four of the six ancestry groups. For the cross ancestry meta analysis, cohort/population with the participants more than 10,000 (European from UK Biobank and European, African, and Latino/Admixed American from All of Us) were used. We performed the meta analysis using MR-MEGA, which accounts for ancestry heterogeneity through genetic principal components.^42^ For the meta-analysis restricted to Europeans (UK Biobank + All of Us), METAL^41^ was used with an inverse-variance weighted fixed-effects model. The visualization and the selection of lead SNPs were performed using gwaslab.^89^ The annotation of lead SNPs was performed with VEP,^90^ OpenTable^91^ and ImmuNexUT platform.^92^

### LD Score Regression and Tissue-specific Heritability Analyses

We performed LDSC analyses to estimate SNP-based heritability and partition it across tissue-and =cell-type–specific annotations.^43,49,53^ GWAS summary statistics were derived from a meta-analysis of the All of Us and UK Biobank European ancestry cohorts, combined using METAL as described above. To ensure consistency, only HapMap3 (HM3) variants were retained during harmonization. Reference LD scores and regression weights for European populations were obtained from the Broad Institute’s public Google Cloud repository (https://console.cloud.google.com/storage/browser/broad-alkesgroup-public-requester-pays). Baseline model v1.2 annotations were used to account for known genomic features.^49^ For each trait, we estimated SNP heritability and genetic correlations using LDSC. Tissue-and cell-type–specific heritability enrichment was assessed using LDSC (--h2-cts) with Multi_tissue_gene_expr annotations.^53^

### Pathway analysis

First, we performed SNP annotation with gene locations (NCBI37.3, https://ctg.cncr.nl/software/MAGMA/aux_files/NCBI37.3.zip) and the reference data created from 1000 genomics Phase3 (g1000_eur, https://ctg.cncr.nl/software/MAGMA/ref_data/g1000_eur.zip) using magma --annotate (with the option, window=10,10).^93^ For the pathway-level analyses, we used MAGMA v1.10 on the same European-ancestry METAL meta-analysis summary statistics used for LDSC. SNPs were first annotated to genes using NCBI37.3 gene locations with a ± 10 kb window and the 1000 Genomes Phase 3 European reference panel (g1000_eur) as the linkage disequilibrium (LD) reference, and variants within the extended MHC region (chr 6: 25-34 Mb) were excluded. Gene-based association tests were then performed with MAGMA using SNP-level p values and sample sizes while accounting for LD from g1000_eur, yielding gene-level z statistics. These results were subsequently used for competitive gene-set analyses that were carried out using MSigDB v2025.1 gene sets, including Hallmark, KEGG canonical pathways, and Gene Ontology (biological process, molecular function, and cellular component) collections.

### PheWAS

We conducted analyses in individuals of European ancestry from the All of Us cohort. ICD-9 and ICD-10 codes were harmonized using PhecodeX.^94^ The R package SPAtest was used to perform score tests based on the saddlepoint approximation, and beta coefficients were estimated using Firth’s method. Visualization was performed with matplotlib.

### EBV detection from scRNAseq data

We developed a pipeline, VIRTUS3, to detect EBV transcripts from 10x Genomics scRNA-seq data. First, we performed alignment to the human reference (GRCh38, refdata-gex-GRCh38-2020-A) and cell barcode calling using Cell Ranger count. The resulting barcodes were used as a whitelist for viral quantification. Unmapped reads were extracted from the position-sorted BAM file with samtools view-f 4 and converted back to FASTQ with cellranger bamtofastq. These unmapped reads were then quantified against an EBV reference (NC_007605.1, B95-8 strain, CDS plus EBER1/2) using Salmon/Alevin ^95^ with the barcode whitelist. The pipeline was applied to the 10x 3’ data of PBMC from EBV-HLH, EBV-IM, and healthy individuals.^55^ We compared the number of EBV-positive cells by VIRTUS3 with the reported number in Suzuki *et al*.^55^ In their analysis, EBV detection was performed using Cell Ranger v5.0 with the custom reference made by the human genome (GRCh38) and the genome of the EBV representative strain (NC_007605).

### Polygenic signal enrichment analysis using scDRS and scDRS-spatial

The polygenic signals were calculated using scDRS^54^ and scDRS-spatial.^57^ The gene scores derived from MAGMA were used. For PBMC scRNAseq data and the visualization of spatial transcriptome data, we used imputed gene expression calculated by MAGIC.^96^ For tissue-and region-specific analysis, we used raw counts for scDRS. The validation of the usage of MAGIC is described in our previous publication.^57^ Polygenic enrichment for each cell was calculated using scDRS (v1.0.3) with --n-ctrl 1000. We used the number of detected genes per cell as a covariate. Group-level statistics were summarized using scdrs perform-downstream and visualized with scdrs.util.plot_group_stats.

### HLA typing and single-marker association testing

We followed the protocol described by Sakaue et al.^97^ Briefly, we used the ACAF PGEN call set of European participants from the All of Us cohort. Sample quality control was performed as in the GWAS. Variant filtering was conducted using PLINK2^98^ with the following options: --maf 0.005 --chr 6 --geno 0.1 --hwe 1e-10 --snps-only just-acgt --min-alleles 2 --max-alleles 2. Variants were lifted over to hg19 using CrossMap, and phasing was performed with EAGLE (v2.4.1).^99^ HLA imputation was carried out using Minimac4 (v4.1.6)^100^ with the publicly available 1KGP HLA panel (https://github.com/immunogenomics/HLA_analyses_tutorial). Single-marker association testing was performed with PLINK2 --glm, adjusting for age, sex, PC1, and PC2 as covariates. We also performed conditioning using PLINK2. The visualization of HLA structure was performed using PyMOL.

### ERAP2-dependent peptide motif analysis

To assess the impact of ERAP2 on the immunopeptidome, we analyzed publicly available immunopeptidomics data (PXD024109^52^) generated from EBV-transformed lymphoblastoid cell lines (LCLs) with ERAP2 knockout using CRISPR-Cas9. W6/32 pan–MHC class I immunoprecipitation data were downloaded and used for downstream analysis.

For motif analysis, the first 9 residues from the N-terminus of each peptide were used, and peptides shorter than 9 amino acids were excluded. Amino acid frequencies across all peptides were defined as background. For ERAP2-dependent and ERAP2-sensitive peptide groups, position-specific amino acid frequencies were calculated. Peptides were classified based on log2 fold change (WT/KO), where ERAP2-dependent peptides were defined as log2FC ≥ 1 and ERAP2-sensitive peptides as log2FC ≤ −1. Motifs were visualized using logomaker.^101^

To quantify ERAP2-dependent sequence features, a sequence-based model was trained using the same dataset. Features included peptide length, amino acid composition, and position-specific one-hot encoding with right padding. A Ridge regression model was trained using these features to predict ERAP2-dependent effects, and the resulting model was used to compute an ERAP2 score for each peptide. As candidate EBV peptides presented on MHC class I peptides, the data generated by Nyeo et al.,^35^ using NetMHCpan,^102^ was used. As a human control dataset, HLA class I positive ligands were extracted from the IEDB database.^103^

### siRNA-mediated knockdown and EBV quantification

JY cells were transfected with pooled siRNAs (three independent siRNAs per gene; Integrated DNA Technologies [IDT]; catalog numbers listed in Supplementary Table 20) targeting six candidate genes. The GWAS highlighted genes that are expressed in JY cells and B cells, and that achieved enough knockdown efficacy in the preliminary experiments were selected as the target genes. A non-targeting siRNA was included as a control. Electroporation was performed using a Lonza 4D-Nucleofector system with SG buffer and program DS-137, according to the manufacturer’s instructions. Following nucleofection, cells were incubated at room temperature for 10 min before the addition of pre-warmed medium and subsequent plating.

Cells were harvested at 24, 48, and 72 h post-transfection. Total RNA and genomic DNA were simultaneously extracted using the AllPrep DNA/RNA 96 Kit (QIAGEN, 80311) according to the manufacturer’s instructions.

For transcript analysis, RNA was reverse-transcribed into cDNA, and EBV gene expression was quantified by TaqMan-based qRT-PCR. Pre-designed TaqMan Gene Expression Assays (Thermo Fisher Scientific) were used for host transcripts, whereas EBV-specific primers and probes were synthesized by IDT. Expression levels were normalized to B2M, and relative expression was calculated using the 2^-ΔΔCt^ method. Catalog numbers and sequences of primer and probe are provided in Supplementary Table 20.

### Genotyping using SNP microarray

We genotyped SNPs from individuals collected at Yale University using the Illumina Global Diversity Array. The genotyping was performed at the Keck Microarray Shared Resource core laboratory at Yale University. We performed standard quality control procedures; variant-level filters included genotyping missingness <5%, MAF > 0.05, HWE p > 1×10^-6^. Genetic ancestry was determined with principal component analysis using 1000 Genomes as a reference panel. Samples of European genetic ancestry were imputed using IMPUTE 2, and strand alignment was performed using SHAPEIT. SNPs with INFO > 0.7 and MAF > 0.01 were retained in the final genomic data.

### Genotyping using Whole Genome Sequencing

We performed whole-genome sequencing on genomic DNA at the Yale Center for Genome Analysis. DNA quantity, purity, and integrity were assessed by spectrophotometry, agarose gel electrophoresis, and Qubit fluorometry. Libraries were prepared from 250 ng genomic DNA using the Watchmaker DNA Library Prep Kit with xGen UDI-UMI adapters, quantified by qPCR, and sequenced on the Illumina NovaSeq X Plus platform to generate 151-bp paired-end reads, targeting at least 135 Gb of passing-filter data per sample. Sequencing data were processed using the Sarek pipeline,^104^ which utilizes GATK HaplotypeCaller.^105^

For a subset of samples, WGS-derived genotypes were harmonized to the imputed array genotype dataset and merged with array-based genotypes for sample demultiplexing.

### Mendelian Randomization Analysis

Two-sample Mendelian randomization (MR) was performed to assess potential causal relationships between EBV DNA positivity and autoimmune diseases. Genetic instruments for EBV DNA positivity were derived from the European-ancestry meta-analysis of EBV DNA GWAS. Instruments were selected at genome-wide significance (*p*<5×10^⁻^) and clumped using PLINK-based LD clumping (*r²*<0.0001, *window*=1000kb) with the European 1000 Genomes Phase 3 reference panel. SNPs with significant heterogeneity in the exposure GWAS (HetPVal<0.05) were excluded prior to instrument selection. Outcome GWAS summary statistics were obtained from publicly available sources: MS from the International Multiple Sclerosis Genetics Consortium (IMSGC), RA from the European-ancestry GWAS, SLE from published meta-analyses, and ischemic stroke from the MEGASTROKE consortium (used as a negative control). Harmonization of exposure and outcome data was performed using the genal Python package, with palindromic SNPs retained (action=1). When exposure SNPs were not directly available in outcome datasets, proxy SNPs were identified using LD-based searching (r²≥0.6, ±10kb window).MR analyses were conducted using the inverse-variance weighted (IVW) method with random effects as the primary analysis. Sensitivity analyses included the Weighted Median method and MR-PRESSO for outlier detection and correction. MR-PRESSO^106^ was applied when evidence of pleiotropy or heterogeneity was detected (Cochran’s Q p<0.0001, or discordant IVW and Weighted Median estimates). When MR-PRESSO identified significant outliers, results were reported after outlier removal. Causal estimates are reported as odds ratios with 95% confidence intervals. To assess the contribution of the MHC region, instruments were stratified into MHC-only (chromosome 6: 25–34Mb) and non-MHC variants, and MR analyses were performed separately for each subset. Bidirectional MR was performed to test for potential reverse causation, using genetic instruments for MS, RA, and SLE as exposures and EBV DNA as the outcome.

### Polygenic risk scoring

PRSs were constructed using PRScs^50^, a Bayesian continuous shrinkage method that infers posterior effect sizes by integrating GWAS summary statistics with an external LD reference panel. The final SNP set for each PRS was determined by the intersection of GWAS variants with the reference panel, yielding 729,766 variants for the genome-wide scores. For stratified analyses, MHC-only and no-MHC PRSs were derived by respectively retaining or excluding variants within the HLA region on chromosome 6 (chr6:25–34 Mb, GRCh38).

For validation in the independent Yale cohort, we used GWAS summary statistics from the European-ancestry meta-analysis (All of Us + UK Biobank) for EBV DNA positivity (binary trait) and EBV DNA load (quantitative trait). The Yale cohort comprises European-ancestry individuals at increased risk of MS (relatives of MS patients and those with morphologically suggestive brain MRI lesions; N=94; mean age 40.2±14.7 years; 71% female) with measured EBV DNA load by qPCR. PRS were calculated for each participant, then standardized to mean 0 and SD 1. Participants were grouped into three PRS categories based on cohort-specific percentiles (≤10%, 10–90%, and ≥90%). EBV DNA positivity was modeled using Firth bias-reduced logistic regression with the PRS as either a continuous predictor (per SD increase) or categorical variable (percentile groups), adjusting for age, sex, and the first 10 genotype principal components. Ordinal trend tests across PRS categories were obtained from models including an ordinal coding of the three groups. EBV DNA load was analyzed using linear regression of log10-transformed EBV DNA copies/mL with the same covariate sets.

For the UK Biobank MS association analysis, we used GWAS summary statistics from the European-ancestry subset of All of Us only (to avoid sample overlap) to construct PRSs that were then applied to European-ancestry individuals in the UK Biobank (N=487,181; 2,423 MS cases; 484,758 controls). PRS values were standardized and participants were grouped by percentiles (≤10%, 10–90%, and ≥90%, or quintiles). Associations with MS were assessed using logistic regression adjusted for age, sex, and the first 10 principal components.

### EBV detection by qPCR

Genomic DNA was extracted from whole blood (1-2mL) using the QIAamp DNA Blood Midi kit (Qiagen, 51183), following the manufacturer’s spin-column protocol. Briefly, blood was incubated with a protease and lysis buffer at 70°C for 10 minutes, mixed with 100% ethanol, and loaded onto the spin column. After centrifugation, columns were washed with AW1 and AW2 buffers, and DNA was eluted with 300µL of elution buffer. DNA was stored at −80 °C before analysis.

EBV genomic DNA was quantified by TaqMan qPCR targeting *BamH1W*. Reactions contained DNA (5µL), Fast Advanced qPCR master mix (Applied Biosystems, 444457), primers, and a FAM-labeled probe. qPCR was performed on a StepOnePlus (Applied Biosystems) with the following cycling conditions: 95 °C for 20 s, followed by 40 cycles of 95 °C for 1 s and 60°C for 20 seconds. An EBV DNA standard (ATCC, VR-3247SD: 6.6×10^5^ copies/µL; Lot: 70063234). Ten-fold serial dilutions of the standard (10^5^ to 10^1^ copies) were prepared and run with each qPCR experiment to define assay sensitivity and ensure plate-to-plate consistency. For each run, a standard curve was generated by plotting Ct values against the log-transformed input copy numbers, followed by calculation of the correlation coefficient (R^2^) from the resulting linear regression.

### Meta-analysis of EBV DNA in patient with MS and controls

We searched PubMed from inception to 2025 using predefined keywords (“multiple sclerosis”, “Epstein–Barr virus”, “PCR”, “viral load”). Only peer-reviewed original research articles that examined EBV DNA in peripheral blood from both MS patients and healthy controls using qPCR were included. Because the sensitivity of EBV DNA detection varies across laboratories and assays, we restricted the analysis to studies in which EBV DNA positivity in either group fell within the range of 5–75 percent to minimize the influence of extreme outliers. When raw counts were not reported, the number of EBV-positive subjects was estimated by multiplying the total sample size by the reported percentage and rounding to the nearest integer. Meta-analysis of odds ratios was performed using the metabin function implemented in the meta package in R.

### Single-cell RNA-seq optimized for EBV detection

PBMCs were isolated from whole blood by density-gradient centrifugation using Lymphoprep (STEMCELL, 18061) and cryopreserved in Bambanker Freezing Media (Bulldog-Bio, BB01) for storage in liquid nitrogen until use. PBMCs were thawed rapidly, and B cells were enriched from PBMCs using a pan-B cell isolation kit (STEMCELL, 19554) or B cell isolation kit (STEMCELL, 19054) according to the manufacturer’s protocols. Enriched B cells were counted and adjusted to a final concentration corresponding to a target loading of 20,000 cells.

Tonsil cells were collected by needle aspiration. Collected samples were centrifuged at 500×g for 5 minutes to remove the supernatant, and the cell pellet was resuspended in 1mL PBS. Total and leukocyte counts were determined using trypan blue and methylene blue, respectively. After counting cells were centrifuged at 500×g for 5 minutes, the supernatant was discarded, and the pellet was cryopreserved in Bambanker Freezing Media for storage in liquid nitrogen until use.

To detect the EBV transcript, a custom EBV-specific primer was added to the cell suspension immediately prior to loading onto the Chromium GEM-X Single Cell 5’ v3 Gene Expression platform (10x Genomics). The primer (sequence: 5′-AAG CAG TGG TAT CAA CGC AGA GTA CAA AAC ATG CGG ACC ACC AGC-3′; 10 µM stock, 1 µL added per sample) was synthesized by Integrated DNA Technologies. Libraries were sequenced on an Illumina NovaSeqXP.

### Single-cell RNAseq analysis

Sequenced reads were processed using Cell Ranger (v8.0.1) with pre-built reference refdata-gex-GRCh38-2024-A and refdata-cellranger-vdj-GRCh38-alts-ensembl-7.1.0 downloaded at 10x Genomics’s Website. EBV transcripts were detected by VIRTUS3. For the pooled samples, donor assignment was performed using the Vireo pipeline^107^ in Demuxafy.^108^ For each sample, cell barcodes and the aligned BAM file generated by Cell Ranger were used as input to generate pileups of exonic SNPs. SNP pileup was performed using cellsnp-lite with the 1000 Genomes Project–derived genotype reference panel (GRCh38; MAF>0.01), applying filters of minimum allele count (minCOUNT≥20) and minor allele frequency (minMAF≥0.1). To restrict genotyping to the relevant donors, we subset the donor VCF to the SNPs detected in the sample-level pileup using bcftools. Vireo was run with an expected donor number of 8 and enabled ambient RNA modeling (--callAmbientRNAs). The downstream single-cell analysis was performed using scanpy 1.10.2.^109^ The embedding and cluster assignment was performed using screfmapping pipeline with pre-defined B cell reference.^6^ Briefly, we performed label transfer using CellTypist^110^ with the model Immune_All_Low. B cell population was extracted based on the predicted cluster. Label transfer and embedding was performed by symphonypy with pre-defined B cell reference (the dataset will be published on Figshare).^111^ BCR were analyzed using airrflow and scirpy.^112,113^ DEG analysis was performed using Nebula^114^, and gene set enrichment analysis was performed using ClusterProfiler.^115^ For tonsil data analysis, we used Azimuth^116^ with the tonsil reference dataset.^117^

### BCR analysis

B-cell receptor analyses were performed using an integrated dataset containing GEX and BCR information. The analysis was restricted to cells from donors with at least one EBV-positive cell. EBV-positive cells were defined as those with EBV gene counts>0. BCR repertoire features, including SHM, were obtained using airrflow.^112^ BCR clones were redefined separately for each donor using productive IGH contigs and scoper,^118^ following the distToNearest, findThreshold, and hierarchicalClones workflow. SHM frequency and clonal expansion were evaluated within each cluster_L2 using models adjusted for donor.

## Supplementary Tables

**Supplementary Table 1 PheWAS of Whole-Blood EBV Positivity in AoU European Participants**

**Supplementary Table 2 PheWAS of Whole-Blood EBV DNA Load in AoU European Participants**

**Supplementary Table 3 Number individuals and** λ**_GC_ of individual GWAS**

**Supplementary Table 4 Lead SNPs of cross-ancestry meta-analysis**

**Supplementary Table 5 Statistics of EBV Positivity GWAS top hits in all single GWAS**

**Supplementary Table 6 Statistics of EBV viral load GWAS top hits in all single GWAS**

**Supplementary Table 7 MAGMA Gene-Set Associations with EBV DNA Positivity**

**Supplementary Table 8 MAGMA Gene-Set Associations with EBV DNA Load**

**Supplementary Table 9 Mendelian Randomization analysis with EBV and autoimmune GWAS**

**Supplementary Table 10 Polygenic Risk Score analysis with EBV GWAS and autoimmune diseases**

**Supplementary Table 11 Polygenic Risk Score analysis with EBV GWAS and EBV DNA positivity in Yale cohort.**

**Supplementary Table 12 HLA allele associated with EBV Positivity**

**Supplementary Table 13 HLA allele associated with EBV DNA Load**

**Supplementary Table 14 HLA amino acids associated with EBV Positivity**

**Supplementary Table 15 HLA amino acids associated with EBV DNA Load**

**Supplementary Table 16 meta analysis of EBV DNA Positivity**

**Supplementary Table 17 Sample information for scRNAseq**

**Supplementary Table 18 Differentially expressed genes in EBV–positive cells**

**Supplementary Table 19 pathway enriched in EBV–positive cells**

**Supplementary Table 20 Reagents and primers used for the experiments**

---

**Figure S1.**
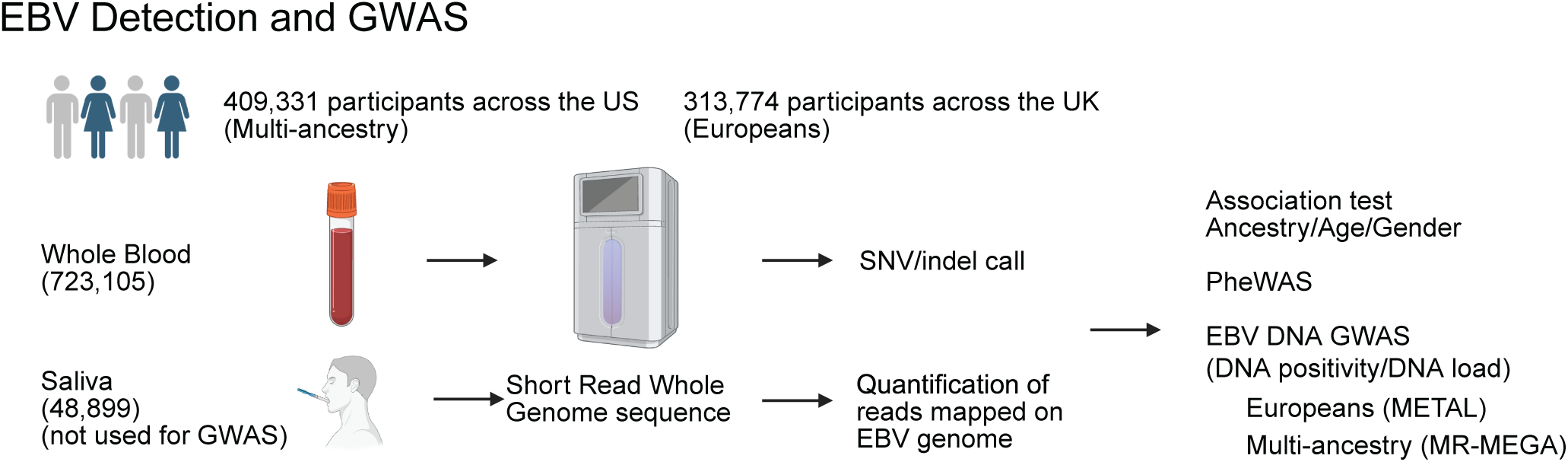
Study design of cross-ancestry meta-analysis of EBV DNA GWAS.

**Figure S2.**
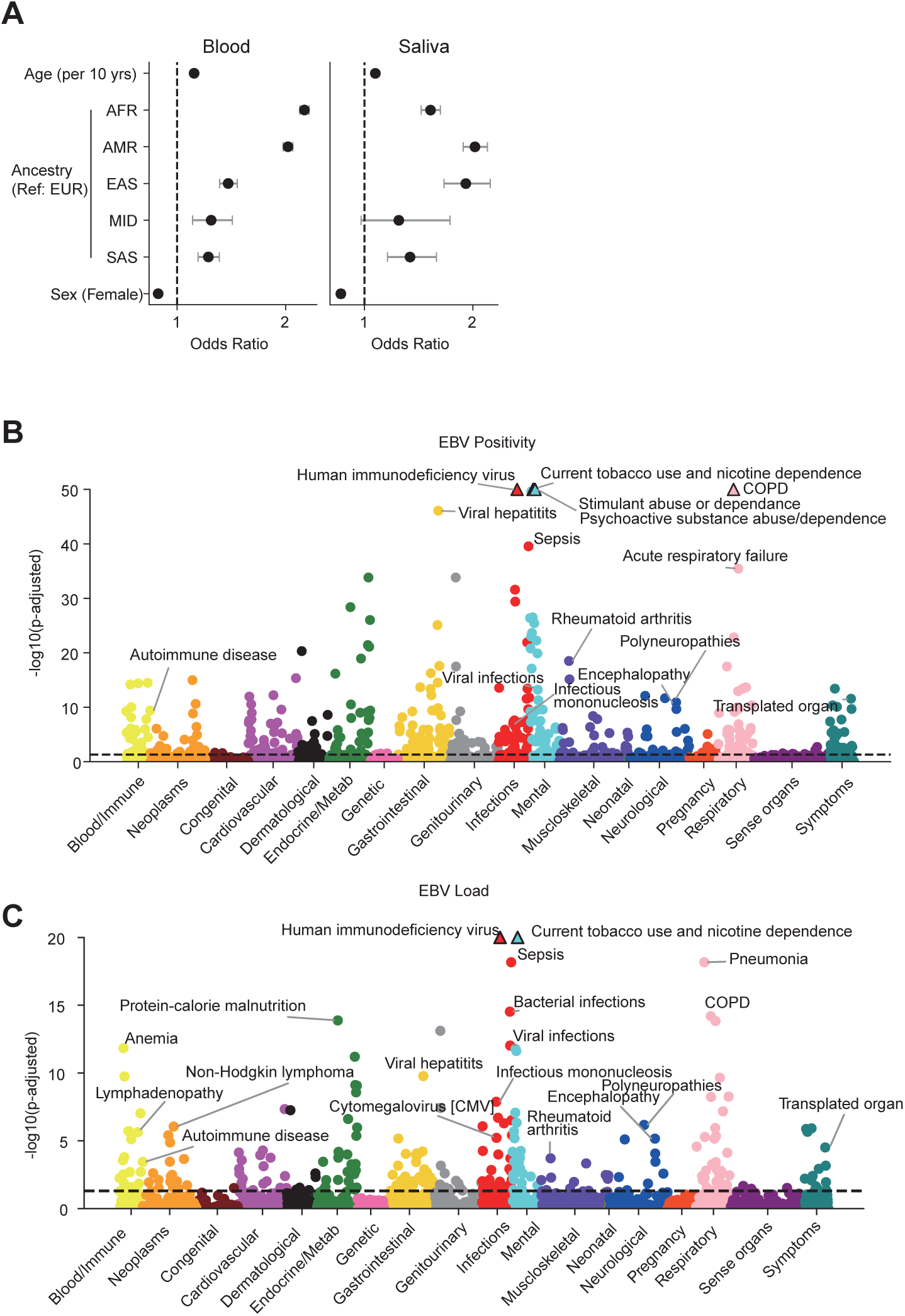
Phenome-wide association study (PheWAS) of DNA EBV viral load in European participants from the All of Us cohort. (A) Forest plots showing odds ratios (points) and 95% confidence intervals (horizontal lines) from logistic regression of EBV DNA positivity for blood samples (left) and saliva samples (right). The age effect represents the change per 10-year increase. European ancestry and male sex were used as references. The dashed line indicates *Odds ratio (OR)*=1. (B and C) Phenome-wide association study (PheWAS) of EBV DNA positivity (B) and EBV DNA load (C) in European participants from the All of Us cohort. Each point represents a clinical phenotype, colored by disease category. The x-axis denotes phenotype groups and the y-axis shows the *–log*₁₀*(FDR-adjusted P value)*. The dashed line indicates the FDR threshold of 0.05. Details are available in Supplementary Tables 1 and 2.

**Figure S3.**
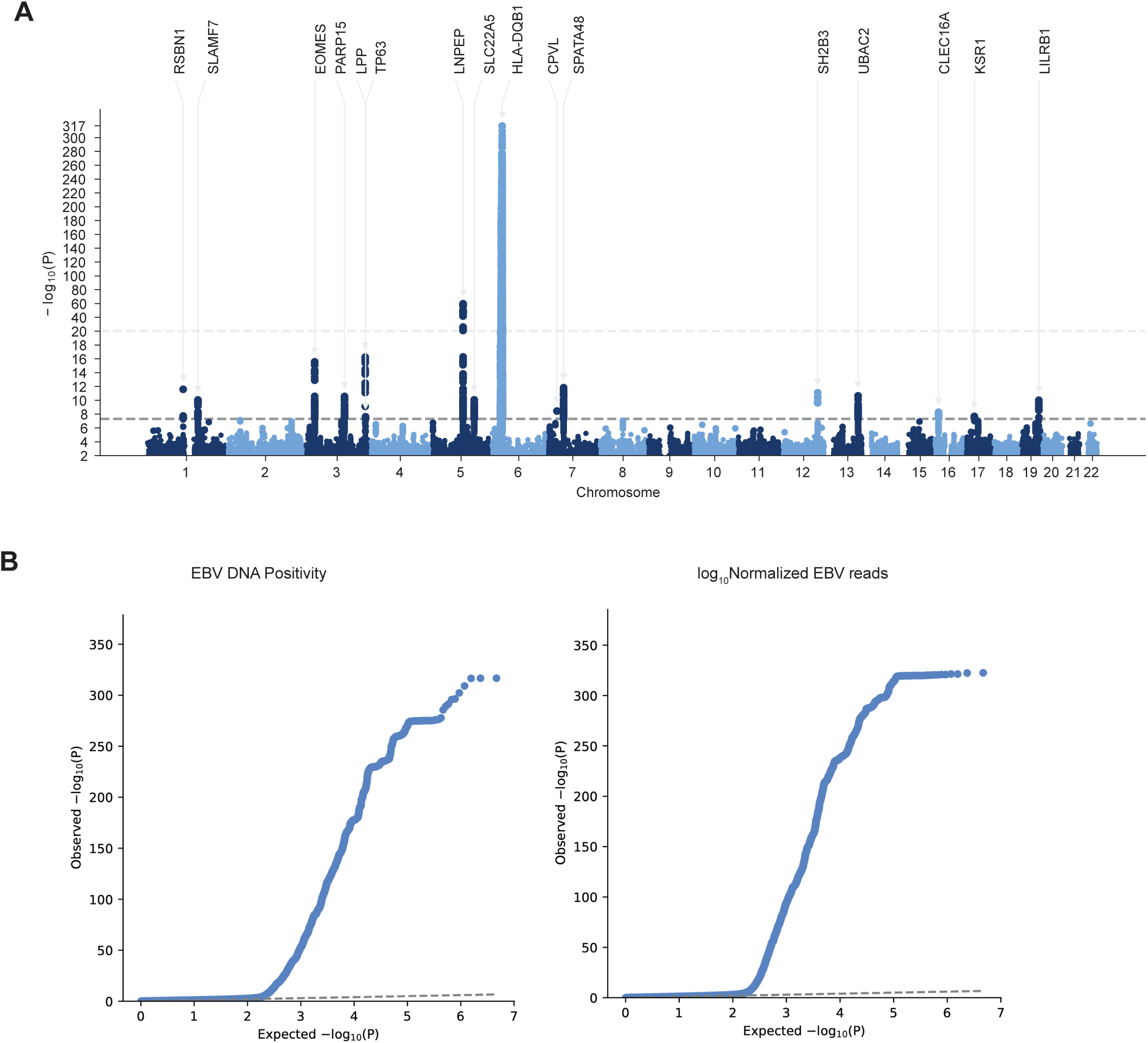
QC plots and annotation of GWAS hits. (A) Manhattan plots showing genome-wide association statistics of EBV DNA load. log_10_(Normalized EBV read) The y-axis shows *-log_10_P* of each variant, and lead SNPs and annotated genes were highlighted. The dashed line indicates a genome-wide significant level (*p*=5.0×10-8). Details are also available in Supplementary Table 4. (B) QQ plot of GWAS for EBV DNA positivity (left) and EBV DNA load (right).

**Figure S4.**
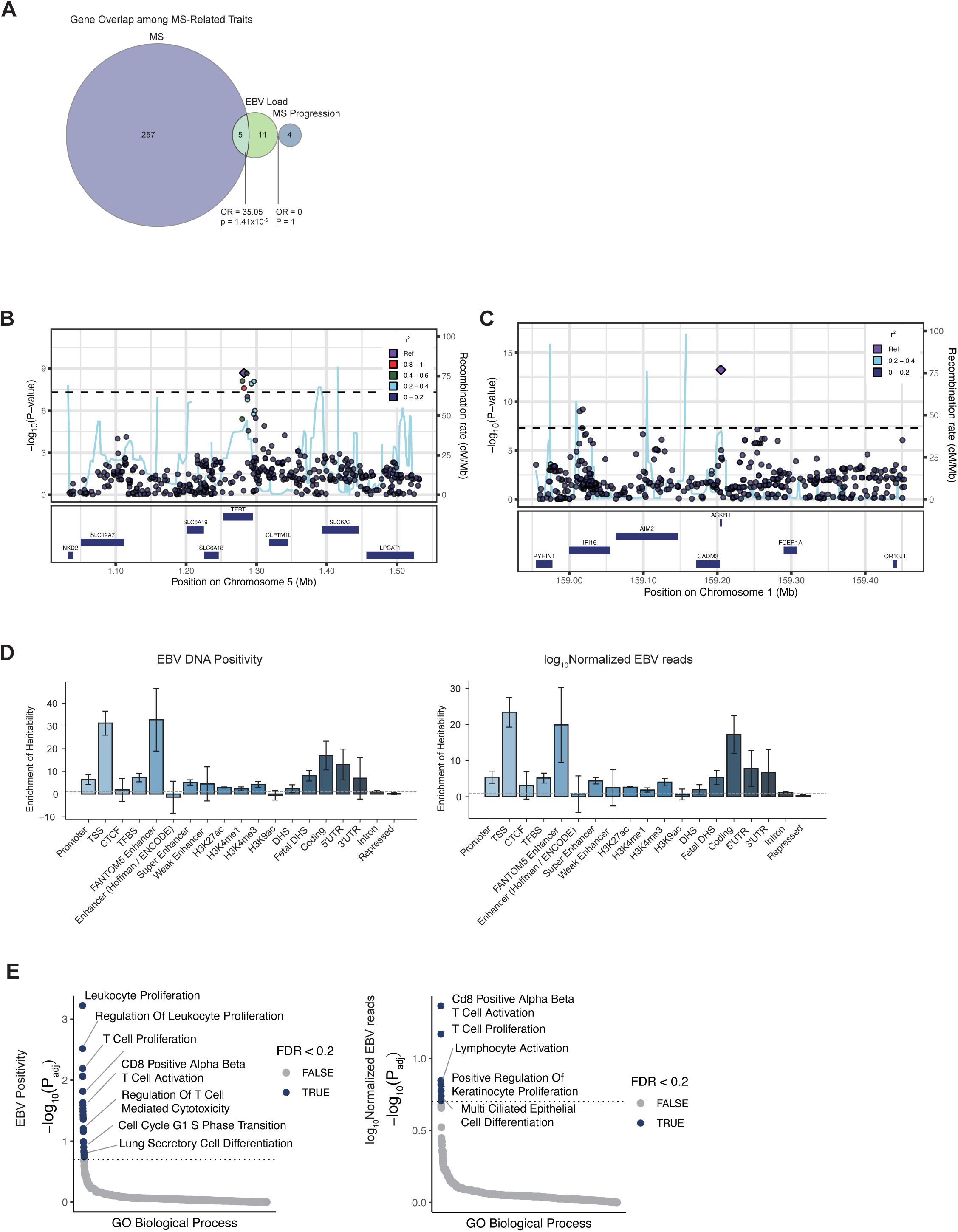
Supportive data for GWAS. (A) Overlap of genes highlighted in MS onset GWAS, MS severity GWAS, and EBV load GWAS. Statistical significance was assessed using a two-tailed Fisher’s exact test. (B and C) Locusplot of rs7726159 (*TERT2*) and rs2814778 (*ACKR1*, located near *IFI16*) in EBV Positivity GWAS (D) Partitioned heritability enrichment in various genomic categories calculated by S-LDSC. EBV DNA positivity (left) and EBV DNA load (right). (E) Gene ontology biological process pathways enriched in GWAS of EBV DNA positivity (left) and EBV DNA Load (right). Each dot represents a pathway, and the size indicates whether the enrichment passed the FDR < 0.2 threshold.

**Figure S5.**
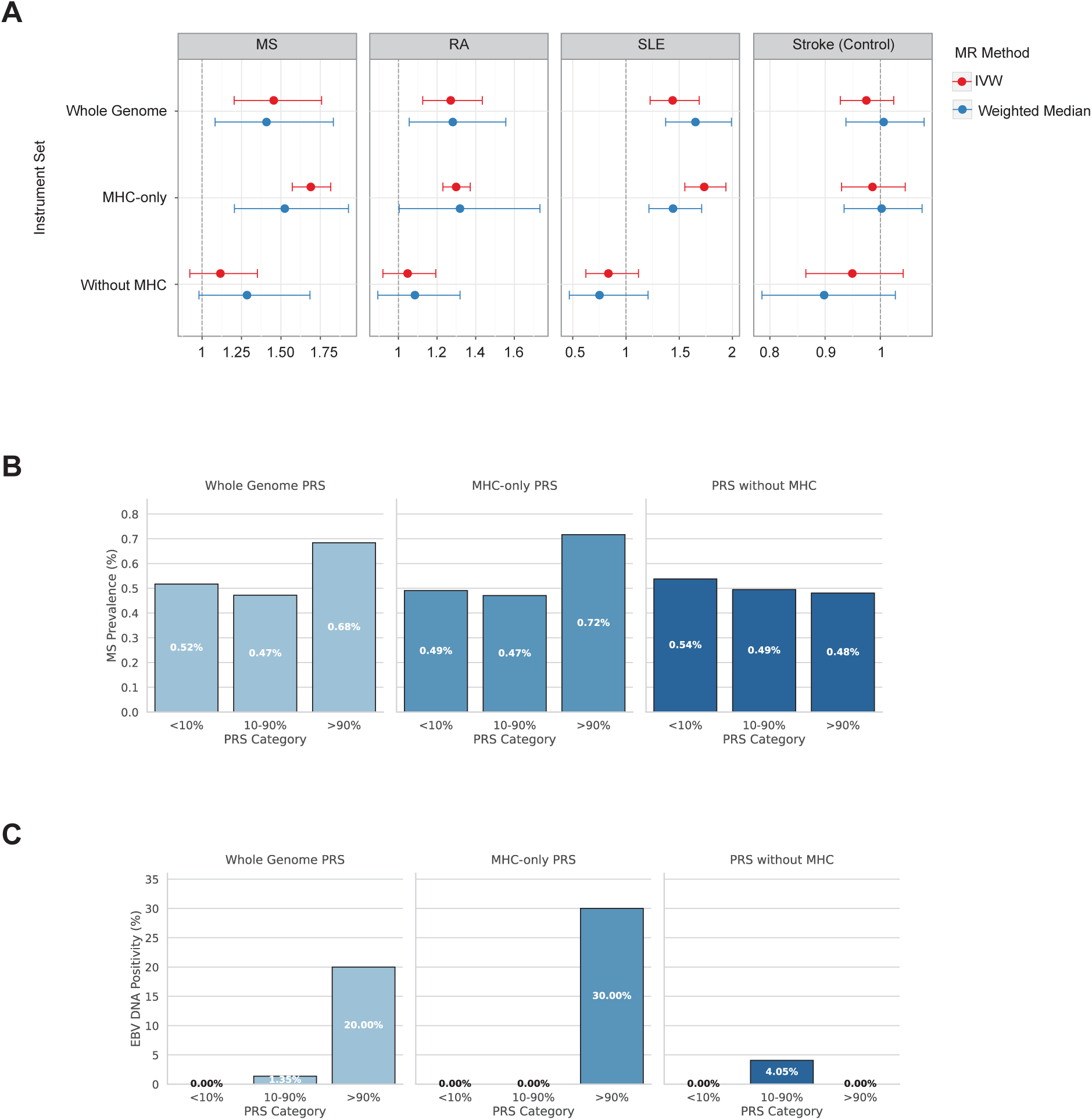
Mendelian randomization and polygenic risk. (A) Forest plots of Mendelian randomization (MR) estimates assessing the causal effect of genetically predicted EBV DNA positivity on autoimmune diseases (multiple sclerosis [MS], rheumatoid arthritis [RA], and systemic lupus erythematosus [SLE]) and stroke (negative control). Three instrument sets were analyzed: whole genome, MHC-only, and genome without the MHC region. Points represent odds ratios; horizontal lines indicate 95% confidence intervals. The dashed vertical line indicates the null effect (OR = 1). (B) MS prevalence across EBV DNA positivity PRS categories in the UK Biobank European-ancestry cohort (N = 487,181; 2,423 MS cases). (C) EBV DNA positivity measured by qPCR across categories of the EBV DNA positivity PRS in an independent Yale cohort of European-ancestry individuals without MS (N = 94; 3 EBV DNA+ cases). For panels B and C, PRSs were constructed using PRScs with All of Us European-ancestry GWAS (B) or the European-ancestry meta-analysis GWAS (C) for three variant sets: whole genome, MHC-only, and non-MHC. Individuals were stratified into bottom 10%, middle 80%, and top 10% of each PRS distribution. Percentages indicate the proportion of MS cases (B) or EBV DNA-positive individuals (C) or within each category.

**Figure S6.**
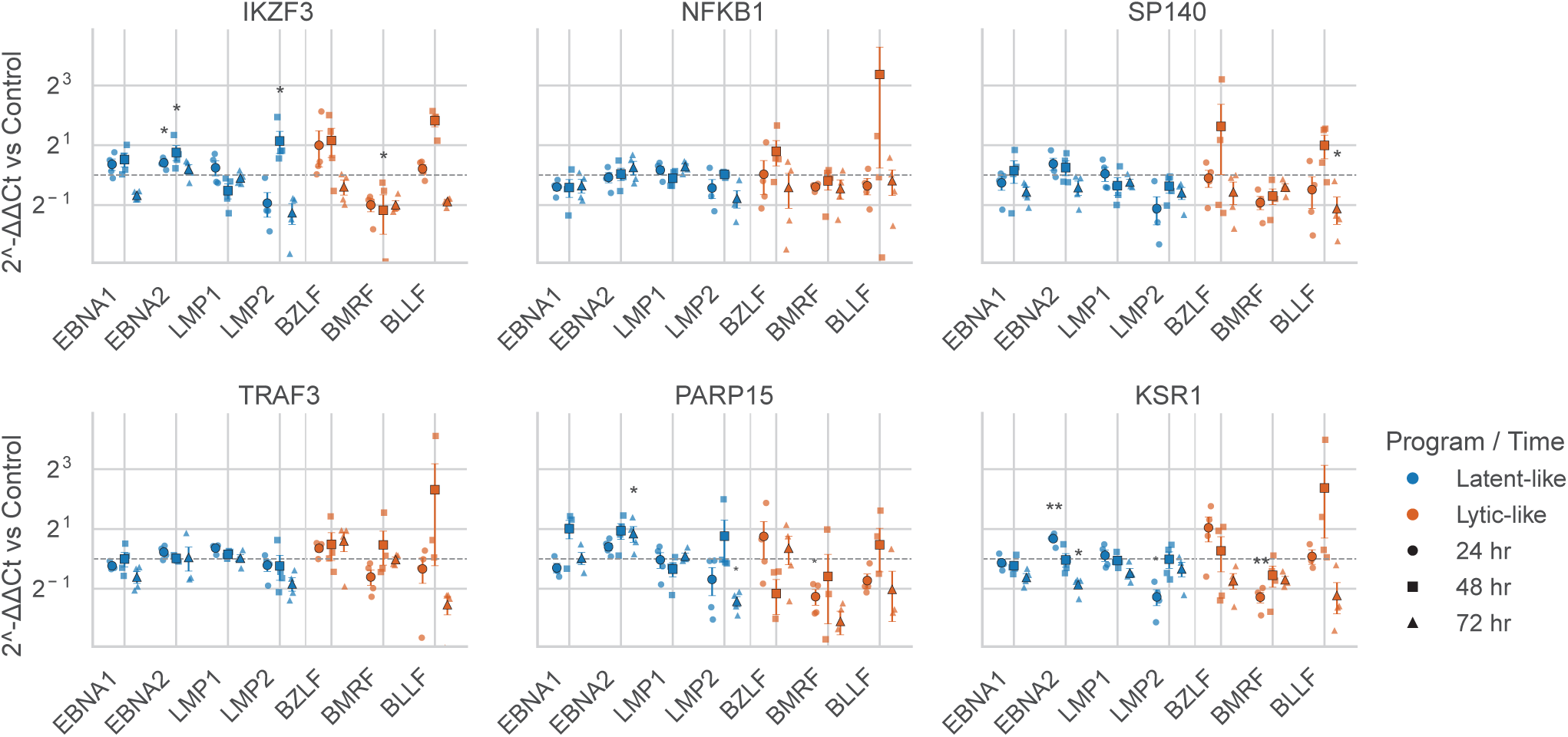
HLA association with EBV DNA load. (A) Locus plot of rs27300 in EBV viral load GWAS. (B) Associations between HLA alleles and EBV DNA load. The size of each point corresponds to the allele frequency, and the top associated alleles are labeled. The alleles highlighted by conditioning were also shown. (C) Sequence logos of ERAP2-dependent (top) and ERAP2-sensitive (bottom) peptides derived from pan–MHC class I (W6/32) immunopeptidomics in EBV LCLs.^52^ Logos represent position-specific enrichment (log2 scale) relative to background amino acid frequencies across all peptides. Positions are indexed from the peptide N-terminus. (D) Distribution of W6/32-derived ERAP2 scores stratified by HLA locus. Top panels show histograms for HLA-A, HLA-B, and HLA-C. Blue indicates human class I ligands from IEDB^103^; orange indicates EBV-derived class I peptides.^35^ Vertical lines denote a score of 0. Bottom panels list the top five EBV peptides for each locus, with corresponding source gene and ERAP2 score.

**Figure S7.**
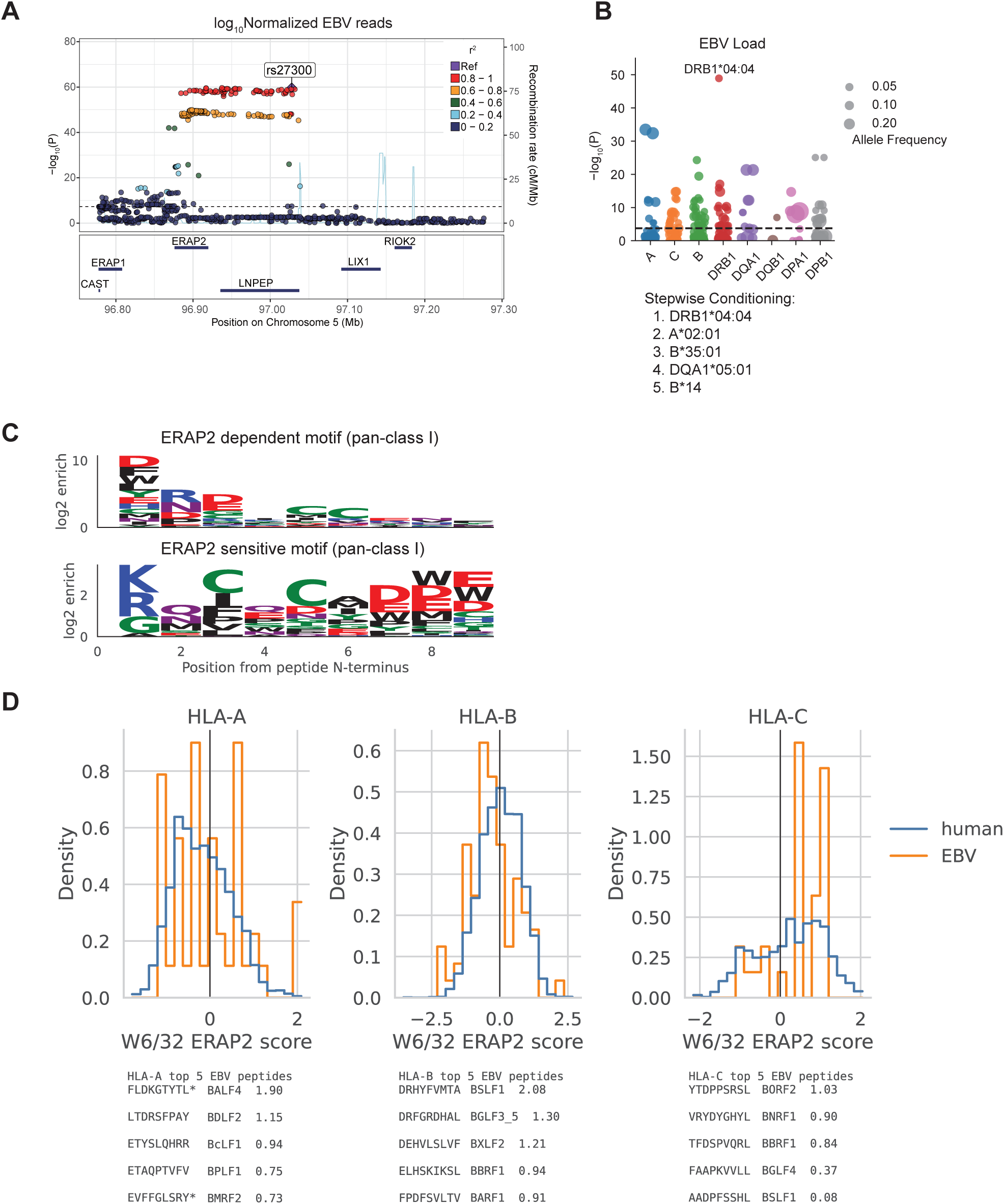
Conditioning of HLA allele and amino acids association with EBV DNA Positivity. Association results for classical HLA alleles across loci (left) and amino acids (right) shown as −log_10_(P values). The most strongly associated alleles in each conditioning round are labeled. HLA amino acid association results for EBV DNA positivity across iterative conditional analyses (Rounds 0–5).

**Figure S8.**
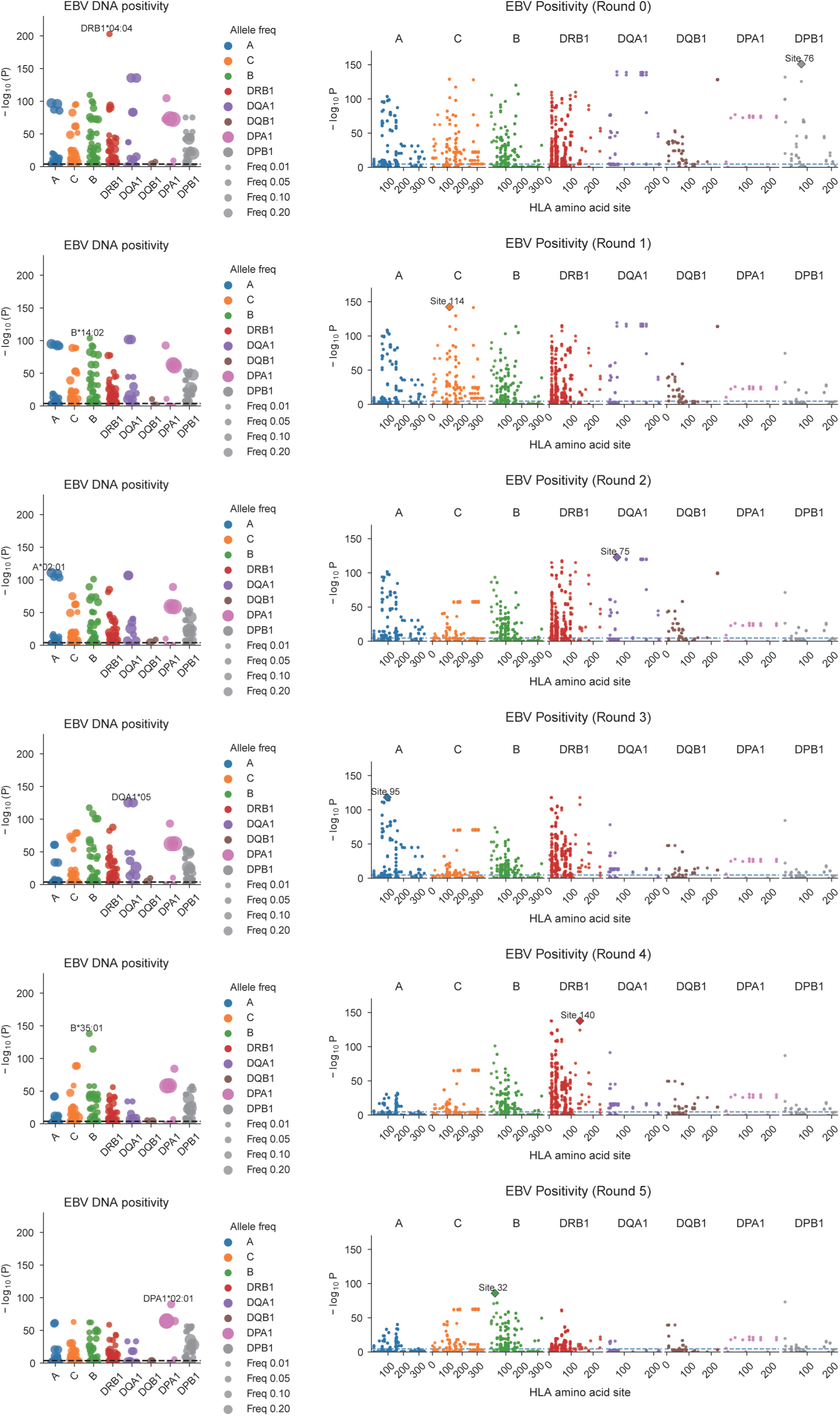
Conditioning of HLA allele and amino acids association with EBV DNA load. Association results for classical HLA alleles across loci (left) and amino acids (right) shown as −log_10_(P values). The most strongly associated alleles in each conditioning round are labeled. HLA amino acid association results for EBV DNA load across iterative conditional analyses (Rounds 0–5).

**Figure S9.**
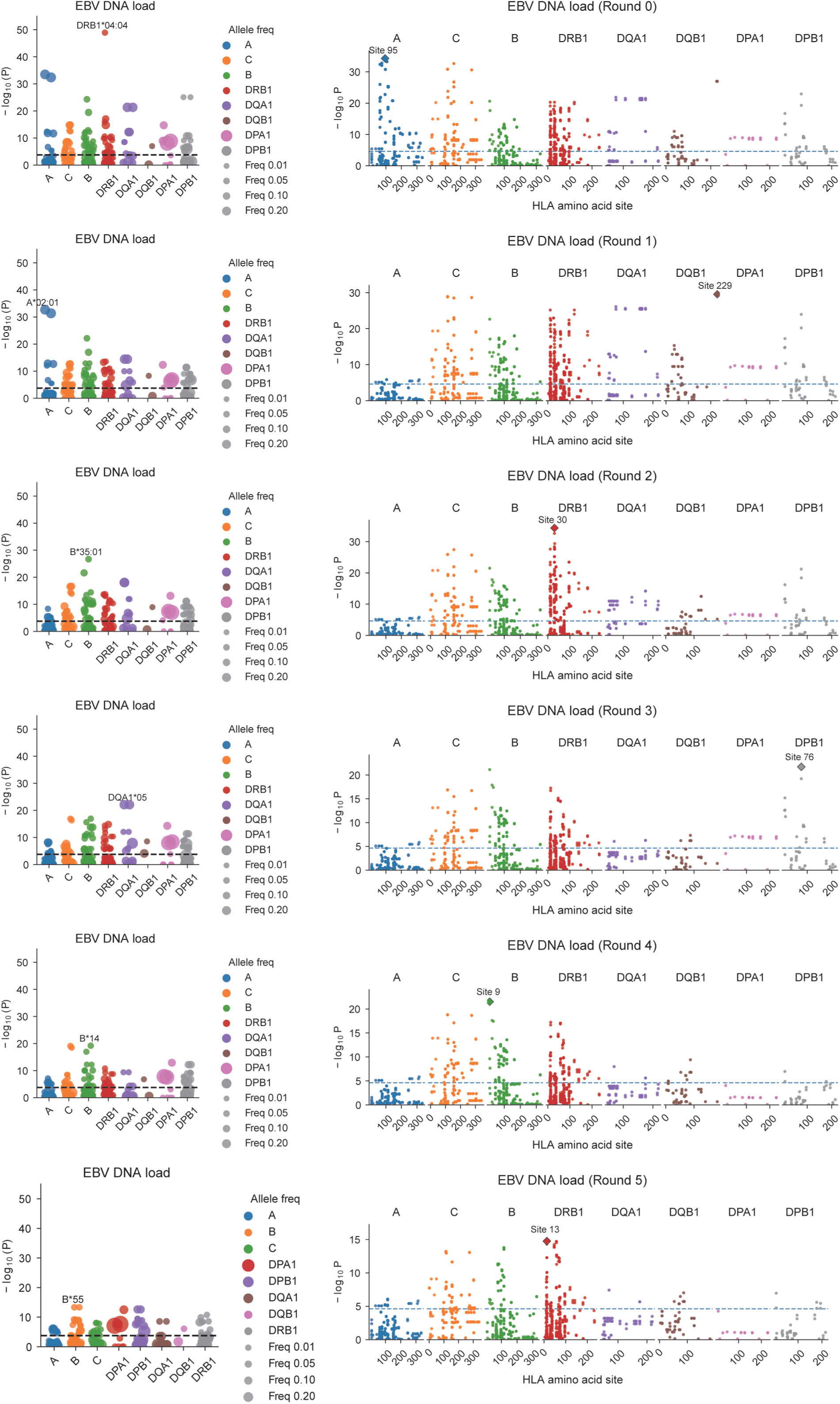
Biological interpretation of GWAS results. (A) Partitioned heritability was estimated using LD score regression for EBV DNA Positivity (left) and EBV DNA load (right). Each dot represents a tissue or cell-type annotation, and the size indicates whether the enrichment passed the *FDR*<0.2 threshold. (A) Proportion of scDRS-significant cells (*FDR*<0.2) across immune cell types for EBV DNA positivity and viral load. (B) Polygenic scores derived from the EBV load GWAS in B memory and plasmablast populations. The Mann-Whitney U test was applied. (C and D) Enrichment in tissues (C) and anatomical regions in tonsils (D) of EBV polygenic signals estimated by scDRS-spatial. Heatmap colors depict the proportion of significant cells (*FDR*<0.2) evaluated using scDRS. Squares denote significant disease associations (*FDR*<0.05), and cross symbols denote significant heterogeneity in association (*FDR*<0.05).

**Figure S10.**
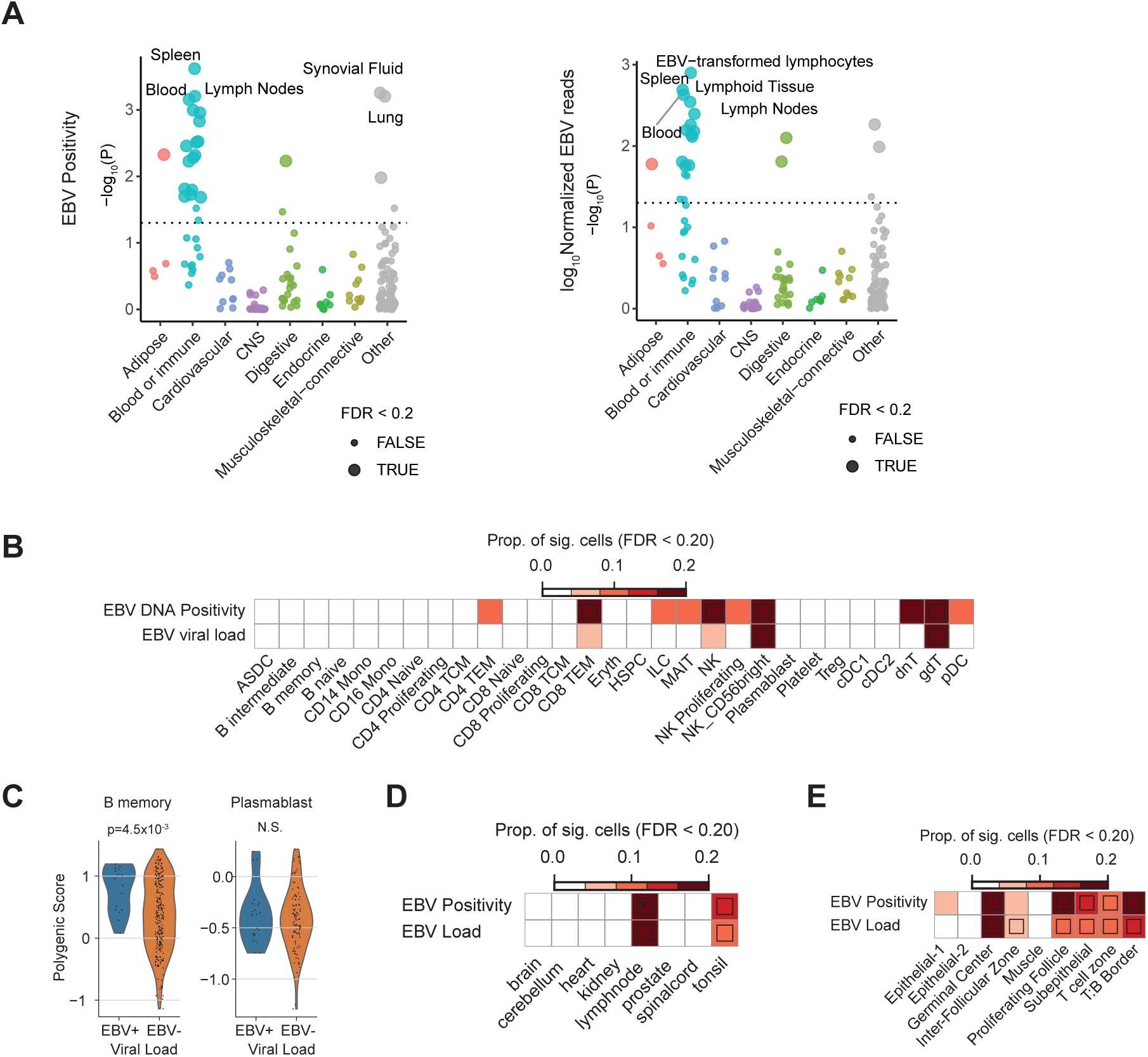
Schematic overview of the VIRTUS3 algorithm for EBV transcript detection in 10x single-cell RNA-seq data. In conventional Cell Ranger-based pipelines using a combined human and EBV reference, overlapping reads can be discarded and reads with sequence similarity to human transcripts can be misaligned, reducing sensitivity for the compact EBV genome. VIRTUS3 addresses this problem through a two-step strategy: reads are first aligned to the human reference with Cell Ranger to generate the human gene-count matrix, and unmapped reads are subsequently recovered and re-analyzed against the EBV transcriptome with Alevin.^95^ This design enables stringent filtering of human-like reads and appropriate handling of multi-mapped viral reads.

**Figure S11.**
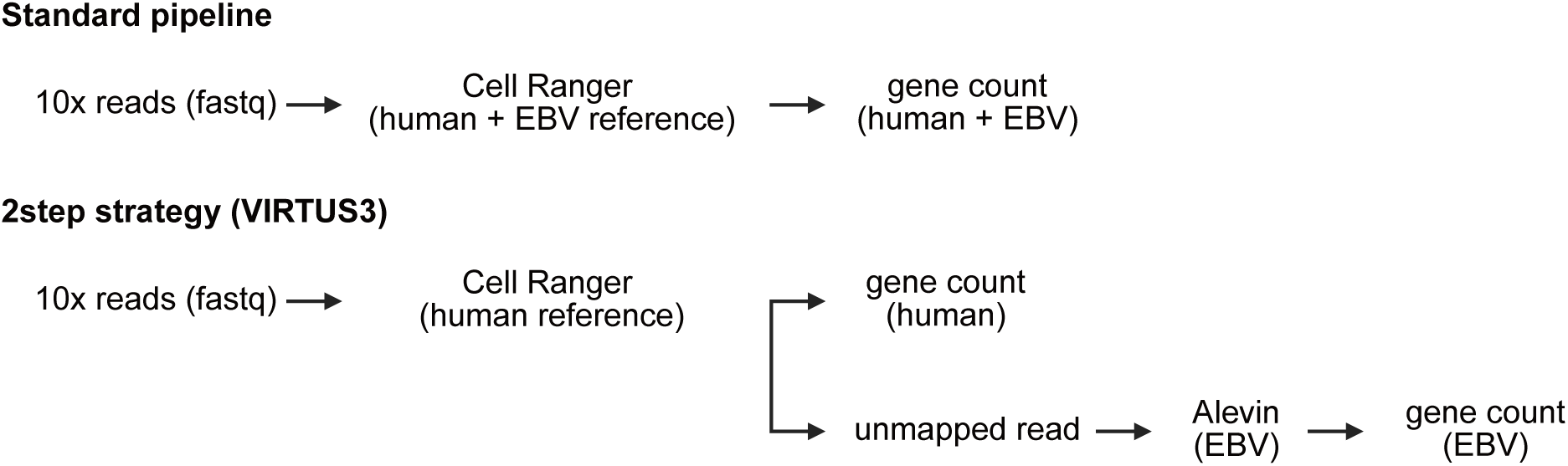
Funnel plot assessing publication bias in studies comparing EBV DNA positivity between MS and controls. Each point represents an individual study included in the meta-analysis. The vertical dashed line indicates the pooled odds ratio under the common-effect model (*OR*=1.77). The dashed diagonal lines denote the 95 percent confidence limits of the expected distribution in the absence of publication bias.

**Figure S12.**
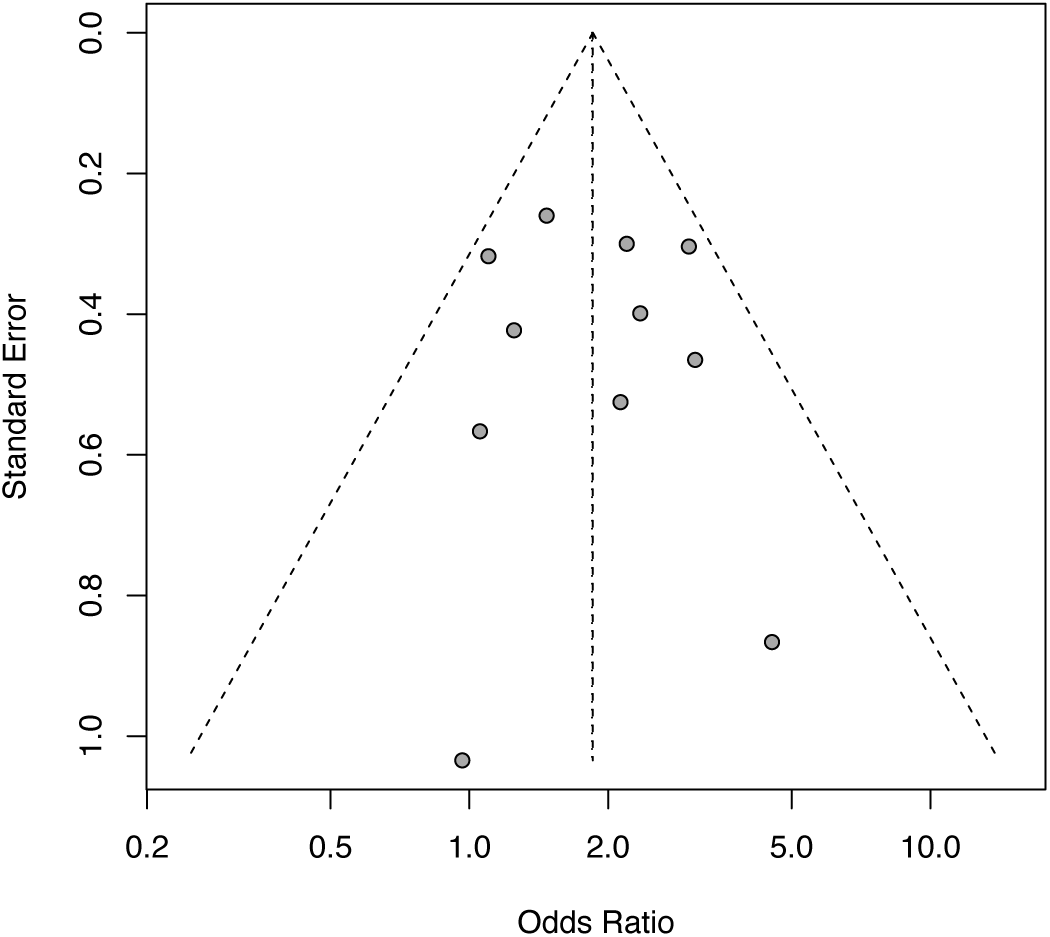
EBV profiling of tonsils from MS and nonMS individuals (A and B) UMAP plot showing predicted clusters (A) and detected EBV–positive cells (B).

**Figure S13.**
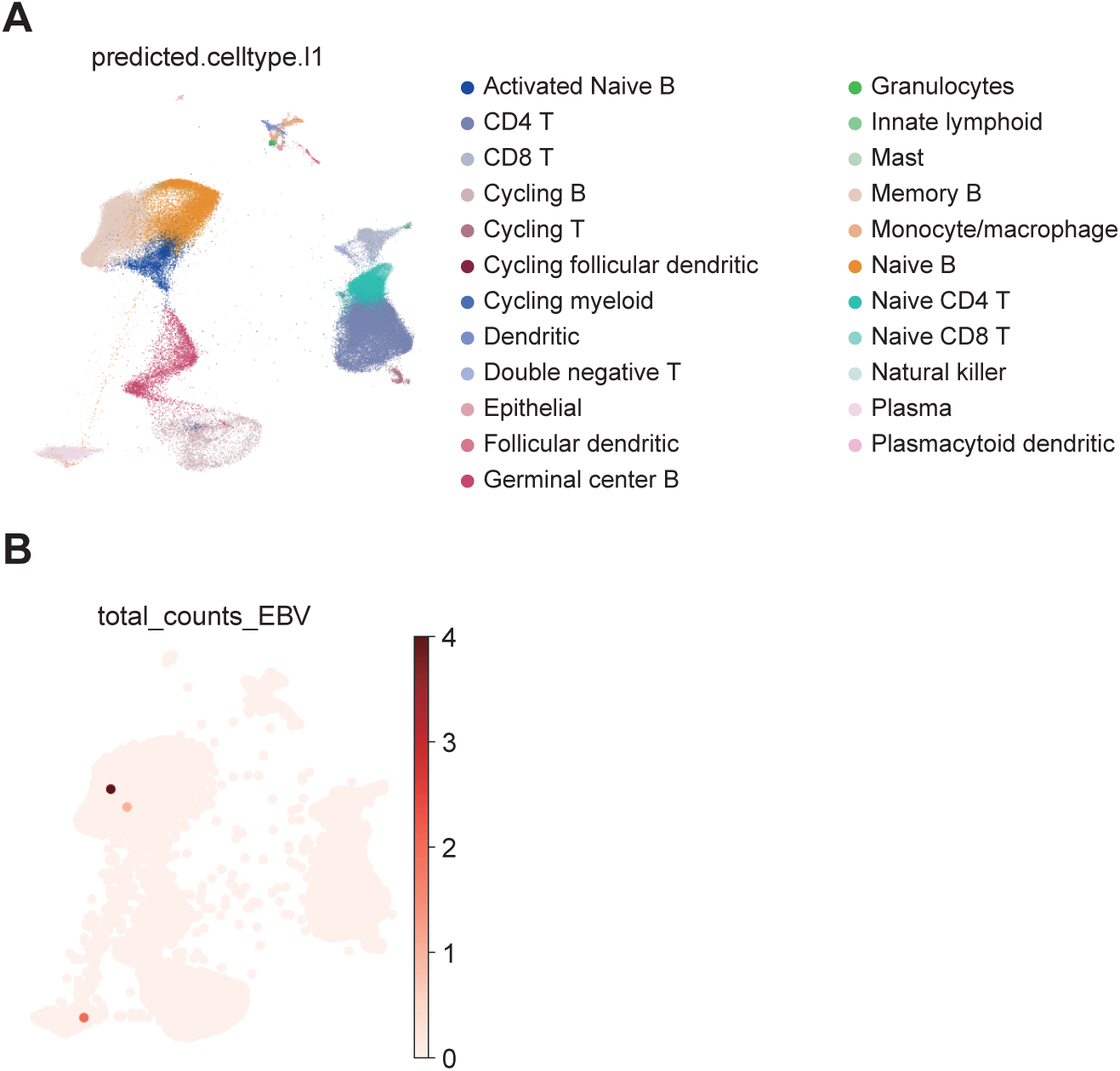
EBV profiling of blood B cells from MS and nonMS individuals (A) Percentage of EBV–positive cells across B cell populations per samples. Rows indicate each donor. The heatmap in orange shows qPCR based copy number of EBV DNA in blood. (B) Forest plot showing the association of isotype with EBV DNA positivity. Clusters were treated as a covariate to extract global isotype preference. IGHE, IGHG3, IGHG4 were removed from the plot because no EBV–positive cells were detected. (C) Somatic hypermutation frequency distribution across clusters between EBV–positive and EBV-negative cells.

**Figure S14.**
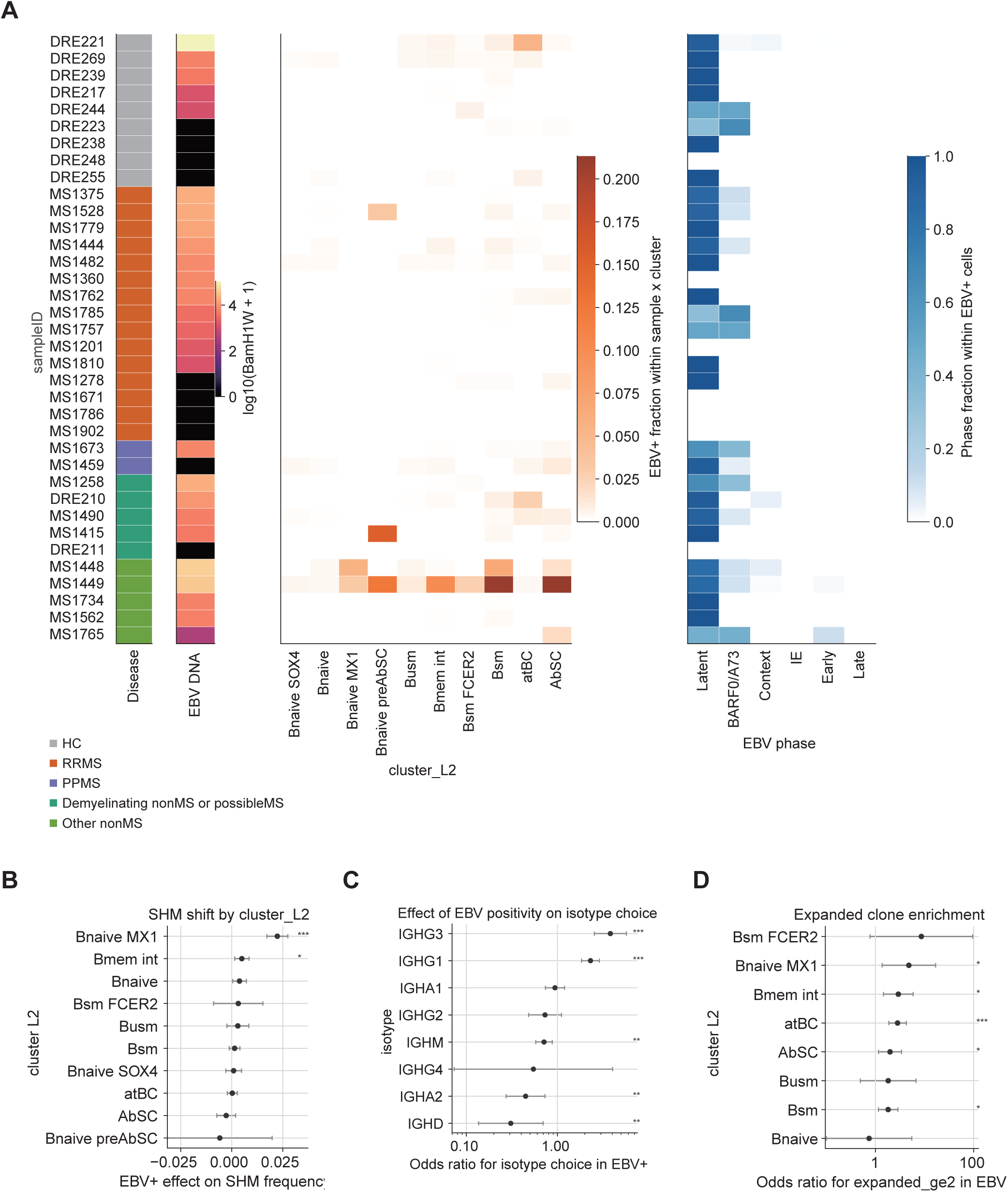
EBV transcriptional states and B-cell receptor features of EBV-positive B cells across donors. (A) Donor-level overview of EBV-positive B cells. Rows represent individual donors. Left annotation bars indicate disease category and whole-blood EBV DNA load measured by qPCR targeting BamH1W. The middle heatmap shows the fraction of EBV-positive cells within each donor-by-cluster_L2 combination. The right heatmap shows the distribution of EBV transcriptional states among EBV-positive cells from each donor, including Latent, BARF0/A73, Context dependent, IE, Early, and Late states. (B) Effect of EBV positivity on somatic hypermutation (SHM) frequency across B-cell subsets. Points indicate effect estimates, and horizontal lines indicate 95% confidence intervals. (C) Association between EBV positivity and immunoglobulin isotype usage. Points indicate odds ratios for isotype choice in EBV-positive cells, and horizontal lines indicate 95% confidence intervals. (D) Enrichment of clonal expansion in EBV-positive cells across B-cell subsets. Points indicate odds ratios for membership in expanded clones (clone size ≥2) in EBV-positive cells, and horizontal lines indicate 95% confidence intervals. Asterisks denote statistically significant associations.

